# Association between inflammation and cognition: triangulation of evidence using a population-based cohort and Mendelian randomization analyses

**DOI:** 10.1101/2022.08.15.22278773

**Authors:** Chloe Slaney, Hannah M. Sallis, Hannah J. Jones, Christina Dardani, CHARGE Inflammation Working Group, Kate Tilling, Marcus R. Munafò, George Davey Smith, Liam Mahedy, Golam M. Khandaker

**Affiliations:** MRC Integrative Epidemiology Unit at the University of Bristol, UK; School of Psychological Science, University of Bristol, 12a Priory Road, Bristol, UK; Centre for Academic Mental Health, Population Health Sciences, Bristol Medical School, University of Bristol, UK; Population Health Sciences, Bristol Medical School, University of Bristol, Bristol, UK; National Institute for Health Research Biomedical Research Centre at the University Hospitals Bristol NHS Foundation Trust and the University of Bristol, UK

**Keywords:** cognition, inflammation, Mendelian randomization, observational, ALSPAC, CRP, IL-6, GlycA, emotion recognition, working memory, response inhibition

## Abstract

**Background:** There is evidence for an association of inflammation with cognitive functioning and dementia in older adults, but the association with cognitive functioning in youth and whether this is causal remains unclear.

**Methods:** In a population-based cohort (Avon Longitudinal Study of Parents and Children; ALSPAC), we investigated cross-sectional associations of inflammatory markers (C-reactive protein [CRP] and Glycoprotein acetyls [GlycA]) with measures of cold (working memory, response inhibition) and hot (emotion recognition) cognition at age 24 (N=3,305 in multiple imputation models). Furthermore, we conducted one-sample and two-sample bidirectional Mendelian randomization (MR) analyses to examine potential causal effects of genetically-proxied inflammatory markers (CRP, GlycA, Interleukin-6, soluble Interleukin-6 receptor) on cognitive measures (above) and general cognitive ability.

**Results:** In the ALSPAC cohort, there was limited evidence of an association between inflammatory markers and cognitive measures at age 24 after adjusting for potential confounders (N=3,305; beta range, -0.02 [95% confidence interval (CI) -0.06 to 0.02, *p*=.29] to 0.02 [95% CI -0.02 to 0.05, *p*=.38]). Similarly, primary MR analyses found limited evidence of potential effects of genetically-proxied inflammatory markers on working memory, emotion recognition or response inhibition in one-sample MR using ALSPAC data (beta range, -0.73 [95% CI -2.47 to 1.01, *p*=.41] to 0.21 [95% CI -1.42 to 1.84, *p*=.80]; or on general cognitive ability in two-sample MR using the latest Genome-Wide Association Study (GWAS) datasets (beta range, -0.02 [95% CI -0.05 to 0.01, *p*=.12] to 0.03 [95% CI -0.01 to 0.07, *p*=.19].

**Conclusions:** Our findings do not provide strong evidence of a potential causal effect of inflammatory markers (CRP, Interleukin-6, GlycA) on the cognitive functions examined here. Given the large confidence intervals in the one-sample MR, larger GWAS of specific cognitive measures are needed to enable well-powered MR analyses to investigate whether inflammation causally influences hot/cold cognition.

## 1. Introduction

Cognitive function predicts many important life outcomes including educational attainment (Deary et al., 2007; Strenze, 2007), occupation status (Schmidt & Hunter, 2004; Strenze, 2007) health-related mortality (Calvin et al., 2017), and quality of life (Cumming et al., 2014). Poorer cognitive functioning is a core feature of many mental health disorders including depression, schizophrenia and Alzheimer’s disease (Bolt et al., 2019; Dalili et al., 2015; Fusar-Poli et al., 2012; Nikolin et al., 2021; Rock et al., 2014), and is highly prevalent in some physical illnesses including cancer (Janelsins et al., 2014; Van Dyk & Ganz, 2021). Despite this, there are no available treatments which effectively address poorer cognitive functioning (Conradi et al., 2011; Hasselbalch et al., 2011). Instead, some treatments may result in worse cognition (Van Dyk & Ganz, 2021). Therefore, there is a need to identify modifiable risk factors that may be targets for prevention and treatment of cognitive impairments.

One promising intervention target could be inflammation (Khandaker et al., 2018). There is some evidence that systemic inflammatory markers such as C-reactive protein (CRP) are associated with poorer cognitive functioning in older adults and in people with physical or mental health conditions (Misiak et al., 2018; Morrens et al., 2022; Sartori et al., 2012). Observational studies in the general population have also reported associations between inflammatory markers, such as CRP, Interleukin-6 (IL-6) and Glycoprotein acetyls (GlycA), and poorer general and domain-specific cognition (Conole et al., 2021; Kokosi et al., 2021; Mac Giollabhui et al., 2021; Shields et al., 2021; van der Lee et al., 2018); although improved cognition has also been reported (Milton et al., 2021). Regarding the human experimental literature, the effect of acute inflammatory challenges (endotoxin and vaccines) on hot (e.g., emotion recognition) and cold (e.g., attention, memory) cognitive domains have yielded inconsistent findings (Balter et al., 2018; Bollen et al., 2017; Brydon et al., 2008; Handke et al., 2020; Harrison et al., 2014). For example, a systematic review reported conflicting results within cold cognitive domains (attention, executive function, memory), with some studies reporting decreased or enhanced performance after an inflammatory challenge, whilst others did not (Bollen et al., 2017). In contrast, there was more consistent evidence in hot cognitive domains of social and emotion processing, where inflammation reduced performance (Bollen et al., 2017). However, most experimental studies are restricted to males and small sample sizes (N<50).

Whilst there is some evidence of an association between inflammation and cognition, there are gaps in the literature. First, current studies are often restricted to small samples (Bollen et al., 2017). Larger studies in the general population are needed to provide more reliable evidence. Second, few studies have examined the association between inflammation and cognition in youth, instead most studies have examined older adults (Sartori et al., 2012). Third, the direction and causality of association are unknown. Specifically, it is unclear whether inflammation affects cognition or if observed associations are due to residual confounding or reverse causation. Fourth, most studies examined commonly measured markers such as IL-6 and CRP (Conole et al., 2021; Kokosi et al., 2021; Mac Giollabhui et al., 2021; Shields et al., 2021). Whilst these are useful markers of systemic inflammation, their levels vary over time within individuals (Bogaty et al., 2013). Investigating novel inflammatory markers such as GlycA, which are thought to be more stable and better reflect chronic inflammation (Connelly et al., 2017; Otvos et al., 2015; Ritchie et al., 2015), could be more fruitful.

In this study, we first examined associations between inflammatory markers (CRP and GlycA) and cold (working memory and response inhibition) and hot (emotion recognition) cognitive measures within a large population-based cohort (Avon Longitudinal Study of Parents and Children; ALSPAC). To better assess causality, we then used one-sample Mendelian randomization (MR) (Sanderson et al., 2022) to examine associations between inflammatory markers (CRP, IL-6, sIL-6R, GlycA) and the same cognitive domains within ALSPAC. Inflammatory exposures were selected based on their well-studied associations with mental health conditions (CRP, IL-6, sIL-6R). We also included GlycA as it is thought to provide a more stable marker of chronic inflammation (Connelly et al., 2017; Otvos et al., 2015; Ritchie et al., 2015). To increase statistical power, we used two-sample MR to examine potential causal relationships between the same inflammatory markers and general cognitive ability (GCA). Given the possibility that the relationship between inflammation and cognition could be in either direction, we also conducted bidirectional analyses which tests both possibilities. Based on previous research, we hypothesised that higher levels of inflammation would be associated with poorer cognition.

## 2. Materials and methods

This study was pre-registered (Open Science Framework: https://osf.io/892wr/), with deviations justified (Supplementary Table S23). Ethics approval was obtained in original studies.

### 2.1. Cross-sectional analysis in the ALSPAC cohort

#### 2.1.1. Cohort description

ALSPAC is a longitudinal population-based birth cohort which initially recruited 14,541 pregnant women in the Avon area (UK) with expected delivery dates between 1^st^ April 1991 and 31^st^ December 1992 (Boyd et al., 2013; Fraser et al., 2013; Northstone et al., 2019). There were 14,062 live births and 13,988 children who were alive at age one. An attempt was made to increase the original sample size when the eldest children were approximately aged seven. This resulted in a total sample size of 15,454 pregnancies (14,901 alive at 1 year of age) when using data after the age of seven. A vast range of variables are available including data on genetics (Supplementary methods), mental and physical health, and cognition. The study website contains details of the data that is available through a fully searchable data dictionary and variable search tool: www.bristol.ac.uk/alspac/researchers/our-data/. For representativeness of ALSPAC, see: www.bristol.ac.uk/alspac/researchers/cohort-profile/.

#### 2.1.2. Inflammatory Exposures at age 24

High sensitivity CRP and GlycA (mainly a1-acid glycoprotein) were assessed in blood samples collected from participants after fasting for at least 6 hours prior to visit. CRP (mg/l) and GlycA (mmol/l) were quantified using an immunoturbidimetric assay (Roche, UK) and 1D proton (1H) Nuclear Magnetic Resonance spectroscopy-based platform (NR; Nightingale Health, Helsinki, Finland), respectively.

#### 2.1.3. Cognitive Outcomes at age 24

Working memory, the ability to temporarily store and manipulate information, was assessed using the *N-*back task (two-back design) (Kirchner, 1958). On each trial, a number is briefly presented (500 ms) and participants are asked to report whether this number is the same or different from the number presented two trials earlier. This task consists of 48 trials with no feedback (8 trials are matches). Prior to this, there are 12 practice trials with feedback. The primary outcome is discriminability index (*d*’) which provides an overall performance estimate. A higher *d*’ indicates better working memory. Individuals who either did not respond on > 50% trials or had a negative *d’* were removed from analyses (N=78).

Emotion recognition, the ability to identify emotion expressions, was assessed using the Emotion Recognition Task (Penton-Voak et al., 2012). On each trial, a face displaying one of six basic emotions (happy, sad, anger, fear, disgust, or surprise) is briefly presented (200 ms) and then immediately covered up. Following this, participants report which emotion was displayed using the six labels. For each emotion, there are eight levels of intensity. This task consists of 96 trials (each emotion presented 16 times). The primary outcome is hits (i.e., number of emotion expressions correctly identified), with a higher score indicating better emotion recognition.

Response inhibition, the ability to suppress a prepotent response, was assessed using the Stop-Signal Task (Logan et al., 1984). On each trial, a letter (X or O; 1,000 ms) is presented and participants are asked to report which letter was displayed, as quickly as possible. However, on 25% trials a tone is presented after the letter (“stop signal”). Participants are asked to inhibit responding on these trials. The task consists of 256 trials (four blocks of 64 trials). The primary outcome is stop-signal reaction time (SSRT). A lower SSRT indicates better response inhibition.

All exposure and outcomes are continuous measures. For distributions of variables, see Supplementary methods. Of the 15,645 participants in ALSPAC, 3,305 individuals had data on all three cognitive measures (working memory, emotion recognition, response inhibition) at age 24.

#### 2.1.4. Potential Confounders

Potential confounders were chosen based on evidence that these variables may be risk factors for inflammation (O’Connor et al., 2009) and cognition and thus may confound the inflammation-cognition association. Potential confounders included: sex, ethnicity, BMI (age 24), maternal education (Degree, A level, O level, Vocational or CSE), maternal socioeconomic status (SES), smoking status (age 24), alcohol use (age 24) and IQ (age 8). For more details, see Supplementary methods.

#### 2.1.5. Statistical analysis

Data were analysed using *Stata* 16 (StataCorp, 2019) using *mvreg* command. Linear regression models examined the cross-sectional association between inflammation (CRP and GlycA) and cognitive measures at age 24. An unadjusted model was first examined (model 1); followed by models adjusted for sex, ethnicity, and BMI at age 24 (model 2); additionally adjusted for maternal education and SES (model 3); additionally adjusted for smoking and alcohol use at age 24 (model 4); additionally adjusted for IQ at age 8 (model 5). We conducted a sensitivity analysis excluding ALSPAC participants who had CRP ≥ 10 mg/l, an indicator of current infection, at age 24 (N=114). Outcomes and exposures were standardised for direct comparisons (i.e., standardised unit change in cognition per one standard deviation change in the inflammatory exposure). Working memory was not normally distributed and natural log transforming this variable did not correct for this. As such, the untransformed raw variable was used in analyses. For complete case and sensitivity analyses, see Supplementary Tables S3-S6.

##### Dealing with missing data

Given the possibility that data are not missing completely at random (MCAR) in ALSPAC (Taylor et al., 2018), we conducted multiple imputation (MI). The rationale for using MI is (1) to improve power in the complete-cases analysis by imputing covariates and exposures (N range in fully adjusted complete case models: 1,686-1,902), (2) we assume the outcome is missing at random (MAR) given the variables in the analysis model, and (3) we assume every exposure and covariate is MAR given the variables in the imputation model. Participants who had data on all three cognitive outcomes at age 24 were included in the analysis (N=3,305). Exposures (inflammatory markers) and potential confounders were imputed. For each set of imputations, 100 datasets were imputed using chained equations. We included the standardised exposures, standardised outcomes, potential confounders, and auxiliary variables in all models. To increase the plausibility that data are MAR, auxiliary variables were included in the MI (Madley-Dowd et al., 2019; White et al., 2011). These were chosen based on their association with incomplete variables (Supplementary methods). For each exposure, we report multivariate linear regression models with imputed data as our primary analysis. For variables included in the imputation models, see Supplementary Methods and Table S1.

### 2.2. Genetic Mendelian randomization (MR) analysis

MR is a method used to assess causality (Davey Smith & Ebrahim, 2003; Sanderson et al., 2022). Genetic variants strongly associated with an exposure are used as proxies for the exposure; which makes this method less susceptible to reverse causation and confounding (Davey Smith & Ebrahim, 2003). The validity of causal inferences drawn from MR relies on three key assumptions: (1) genetic variants are robustly associated with the exposure, (2) genetic variants are not associated with potential confounders, (3) genetic variants are associated with the outcome only via the exposure. Here, we conducted one-sample MR within the ALSPAC cohort and two-sample MR using publicly available GWAS. Given the possibility that the relationship between inflammation and cognition could be in either direction, we conducted bidirectional analyses to investigate both possibilities. For more details, see Supplementary methods.

#### 2.2.1. One-sample bidirectional Mendelian randomization in the ALSPAC cohort

One-sample MR assessed whether there is a potential causal relationship between the same inflammatory markers and cognitive measures reported in the cross-sectional analysis (N range: 1,677-2,193). An additional inflammatory marker (IL-6 at age 9), not available at age 24, was included.

##### 2.2.1.1. Data Sources

The GWAS listed in Table 1 were used to identify genetic variants (Single Nucleotide Polymorphisms; SNPs) associated with inflammation (CRP, IL-6, sIL6R and GlycA) and cognition (working memory, emotion recognition, response inhibition) (Ahluwalia et al., 2021; Han et al., 2020; Kettunen et al., 2016; Ligthart et al., 2018; Mahedy et al., 2021; Rosa et al., 2019; Sarwar et al., 2012; Swerdlow et al., 2012). For details on GWAS (including any overlap with ALSPAC) and accessing data, see Supplementary Tables S8 and S10. For number of genetic variants included in each SNP set, see Table 1.

**Table 1.** GWAS and instruments used to extract SNPs for inflammation (CRP, IL-6, sIL-6R, GlycA) and cognition (working memory, emotion recognition, and response inhibition).

| Phenotype | GWAS/Instrument | N | SNP location | N | 1SMR | 2SMR |
| --- | --- | --- | --- | --- | --- | --- |
| **Inflammation** |  |  |  |  |  |  |
| *Primary analysis* |  |  |  |  |  |  |
| CRP | Ligthart et al. (2018) | 204,402 | *CRP* gene *cis* SNPs | 6 | 6 | 6 |
|  | Han et al. (2020) | 418,642 | *CRP* gene *cis* SNPs | 20 | 18 | 13 |
| IL-6 | Ahluwalia et al. (2021) | 52,654 | *IL6R* gene *cis* SNPs | 2 | 2 | 2 |
| sIL-6R | Rosa et al. (2019) | 3,301 | *IL6R* gene *cis* SNPs | 34 | 34 | 22 |
| GlycA | Borges et al. (2020) | 115,078 | Genome-wide SNPs | 87 | 82 | 82 |
| *Secondary analysis* |  |  |  |  |  |  |
| CRP | Ligthart et al. (2018) | 204,402 | Genome-wide SNPs | 78 | 76 | 77 |
|  | Han et al. (2020) | 418,642 | Genome-wide SNPs | 552 | 520 | 494 |
| IL-6 | Ahluwalia et al. (2021) | 52,654 | Genome-wide SNPs | 3 | 3 | 3 |
|  | Sarwar et al. (2012) | 27,185 | *IL6R* gene *cis* SNPs | 1 | 1 | 1 |
|  | Swerdlow et al. (2012) | ≤ 4,479/<br>SNP | *IL6R* gene *cis* SNPs | 3 | 3 | 3 |
| GlycA | Kettunen et al. (2016) | 19,270 | Genome-wide SNPs | 10 | 10 | 10 |
| **Cognition** |  |  |  |  |  |  |
| Working memory | Mahedy et al. (2021) | 2,471 | Genome-wide SNPs | 3 | 3 | N/A |
| Emotion recognition | Mahedy et al. (2021) | 2,560 | Genome-wide SNPs | 6 | 6 | N/A |
| Response inhibition | Mahedy et al. (2021) | 2,446 | Genome-wide SNPs | 6 | 6 | N/A |

##### 2.2.1.2. Extracting genetic variants for inflammation and cognition

SNPs were extracted from GWAS full summary statistics based on the following criteria: (1) *p*-value threshold for inflammatory markers (*p*< 5 × 10^−8^) and for cognitive measures (*p*< 5 × 10^−6^; due to no SNPs meeting criteria at 5 × 10^−8^), (2) linkage disequilibrium (LD) clumping (r^2^ =0.01, kb=1000) using *ld_clump()* in the *ieugwasR* package and (3) minor allele frequency > 0.01. LD clumping ensures that SNPs are not highly correlated with each other and are therefore independent. For ease of interpretation, all effect alleles refer to the exposure-increasing allele. Where possible, SNPs were divided into *cis* variants (SNPs +/- 1mB from protein coding gene based on Genome Reference Consortium Human Build 37 or 38, see Supplementary Table S11 footnote for location of protein coding genes) and genome-wide variants (SNPs that met statistical criteria based on *p*-value and LD thresholds). *Cis* variants, due to their proximity to the protein coding gene, are less likely to be pleiotropic (i.e., less likely to influence the outcome via pathways other than the exposure) and therefore may provide more valid instruments. Genome-wide variants may increase statistical power in the analyses due to the larger number available. The primary analysis was performed using *cis* SNPs extracted from the largest available GWAS, except for GlycA which does not have a protein coding gene. Instead, we used the largest GlycA GWAS to date in the primary analysis. Secondary analyses used smaller GWAS/instruments.

##### 2.2.1.3. Creating genetic risk scores for inflammation and cognition

Genetic and phenotypic data were available for 8,130 ALSPAC participants. For one-sample MR, identified SNP sets (Table 1) were combined into a weighted genetic risk score for inflammatory (CRP, IL-6, GlycA, sIL-6R) and cognitive (working memory, emotion recognition, response inhibition) phenotypes for each ALSPAC participant (Purcell et al., 2007). Specifically, risk alleles were weighted by the effect size (beta) reported in the GWAS and then summed to provide a risk score. Unrelated individuals were kept, withdrawals of consent were removed, missing genotypes were not imputed. For SNPs not available in ALSPAC, proxies were identified that had: r^2^ > 0.8 (using *LDproxy_batch* function in EUR population in *R*), rsID available, SNP available in full summary statistics and ALSPAC. For quality checks and number of SNPs included in each genetic risk score, see Table 1, Supplementary Methods, and Table S12.

##### 2.2.1.4. Data analysis

Analyses were carried out in *R* 4.1.1 (R Core Team, 2020). Genetic risk scores were created in Plink v1.90 and two-stage least squares regressions were conducted using the *AER* package (Kleiber & Zeileis, 2008). Regressions examined whether genetic risk scores predicted standard deviation change in the outcome *via* the exposure (IL-6 at age 9, CRP at age 24, GlycA at age 24). Models included top 10 genetic principal components to adjust for genetic ancestry. Outcome variables are standardised for direct comparisons.

##### 2.2.1.5. Two-sample bidirectional Mendelian randomization

Two-sample MR assessed whether there is a potential causal relationship between the same inflammatory markers and general cognitive ability in ∼418,642 individuals.

##### 2.2.1.6. Genetic instruments

For inflammatory markers, we extracted SNPs from the same GWAS/instruments used in the one-sample MR (Table 1). For GCA, we used the largest combined GWAS on GCA to date (Lam et al., 2021). SNPs for GCA were identified using the same criteria outlined in 2.2.1.2, identifying 250 SNPs. Proxies were identified for SNPs not available in the outcome GWAS using the following criteria: R^2^ > 0.8, SNP with highest R^2^ available in both the exposure and outcome summary statistics. For final N SNPs used in each instrument, see Table 1, Supplementary Tables S16 and S19.

##### 2.2.1.7. Additional MR assumption

The same assumptions for one-sample MR apply, with the additional assumption that samples come from similar but non-overlapping populations (Lawlor, 2016). Whilst it is difficult to determine the exact percentage of overlap, there is overlap between most exposure and outcome GWAS due to the use of large data sources (cohorts in CHARGE consortium and UK Biobank). Consequently, some advantages (e.g., weak instruments biasing results towards the null) of two-sample MR may be reduced (Burgess et al., 2016; Lawlor, 2016), although the main advantage of increased statistical power remains (Lawlor, 2016).

##### 2.2.1.8. Data analysis

Two-sample MR was conducted using the *TwoSampleMR* Package 0.5.6 (Hemani et al., 2018) in *R* 4.1.1 (R Core Team, 2020). For each instrument, this package harmonises the SNP-exposure and SNP-outcome data. Palindromic SNPs were excluded if the strand could not be inferred from the minor allele frequency (>0.42), if available. The primary analysis used either the inverse-variance weighted (IVW) method (>1 SNP available) or Wald ratio (1 SNP available). Sensitivity analyses included MR-Egger (Bowden et al., 2015), weighted median (Bowden et al., 2016), weighted mode (Hartwig et al., 2017), and MR Pleiotropy RESidual Sum and Outlier (MR-PRESSO) (Verbanck et al., 2018). As these MR methods have different assumptions regarding the validity of the genetic instruments, we can be more confident in our causal inferences if the relationship between inflammation and cognition is observed across methods. Some MR methods require a minimum number of SNPs, therefore sensitivity analyses are reported when enough SNPs are available. For details on each method, see Supplementary methods. Additional sensitivity analyses included Steiger filtering to check that SNPs have a stronger association with the exposure than the outcome (Hemani et al., 2017), checking for heterogeneity (Cochran’s *Q*-statistic), checking for pleiotropy (Egger intercept).

## 3. Results

Fig. 1 presents an overview of the analyses conducted in this study. For descriptive statistics on cognitive measures, inflammatory markers, and confounders in ALSPAC, see Supplementary Table S2.

**Fig. 1.**
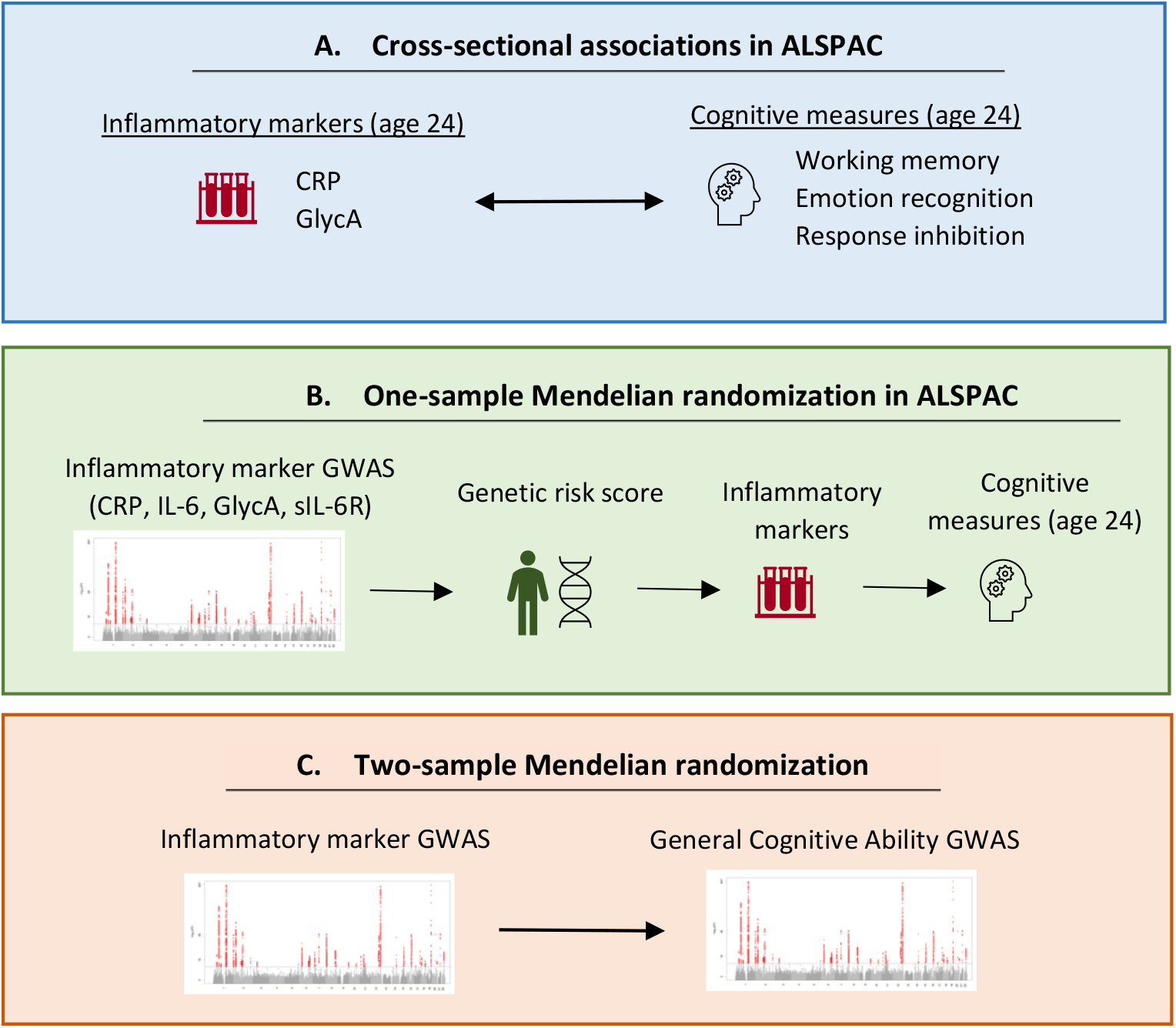
Overview of analyses performed in this study. *Note:* MR was run bidirectionally. Manhattan plot image taken from Ligthart et al. (2018).

### 3.1. Association between inflammatory markers and cognitive function at age 24 in the ALSPAC cohort estimated using multivariate regression

There was limited evidence of an association between CRP and cognitive measures (*p*s ≥ .24; Table 2). GlycA was associated with poorer working memory (β = -0.08, 95% CI = -0.11 to -0.04, *p*< .001), emotion recognition (β= -0.05, 95% CI = -0.09 to -0.01, *p*=.008) and response inhibition at age 24 (β= 0.05, 95% CI = 0.01 to 0.08, *p*=.014) (Table 2), but these associations did not persist after adjusting for potential confounders. This is broadly consistent with complete case analyses and sensitivity analyses (removing individuals with possible infection) (Supplementary Tables S3-S6).

**Table 2.** Cross-sectional associations between inflammatory markers (CRP and GlycA) and cognitive measures at age 24 in ALSPAC (N=3,305; multiply imputed models)

| **Models** | ***b*** | **95% CI** | ***p*** |
| --- | --- | --- | --- |
| **C-reactive Protein (CRP)** |  |  |  |
| *Outcome: Working memory* |  |  |  |
| Model 1 | -.02 | -.06 to .01 | .24 |
| Model 2 | -.005 | -.04 to .03 | .80 |
| Model 3 | -.003 | -.04 to .04 | .88 |
| Model 4 | .002 | -.04 to .04 | .90 |
| Model 5 | .01 | -.03 to .05 | .60 |
| *Outcome: Emotion Recognition* |  |  |  |
| Model 1 | -.01 | -.04 to .03 | .73 |
| Model 2 | .004 | -.03 to .04 | .84 |
| Model 3 | .005 | -.03 to .04 | .79 |
| Model 4 | .009 | -.03 to .05 | .61 |
| Model 5 | .02 | -.02 to .05 | .38 |
| *Outcome: Response Inhibition* |  |  |  |
| Model 1 | .02 | -.02 to .05 | .39 |
| Model 2 | -.004 | -.04 to .03 | .82 |
| Model 3 | -.004 | -.04 to .03 | .83 |
| Model 4 | -.01 | -.05 to .03 | .58 |
| Model 5 | -.02 | -.05 to .02 | .40 |
| **Glycoprotein Acetyls (GlycA)** |  |  |  |
| *Outcome: Working memory* |  |  |  |
| Model 1 | -.08 | -.11 to -.04 | <.001 |
| Model 2 | -.05 | -.09 to -.01 | .022 |
| Model 3 | -.04 | -.08 to .003 | .072 |
| Model 4 | -.03 | -.07 to .01 | .17 |
| Model 5 | -.02 | -.06 to .02 | .29 |
| *Outcome: Emotion recognition* |  |  |  |
| Model 1 | -.05 | -.09 to -.01 | .008 |
| Model 2 | -.01 | -.05 to .03 | .63 |
| Model 3 | .002 | -.04 to .04 | .94 |
| Model 4 | .008 | -.03 to .05 | .68 |
| Model 5 | .01 | -.02 to .05 | .46 |
| *Outcome: Response Inhibition* |  |  |  |
| Model 1 | .05 | .01 to .08 | .014 |
| Model 2 | .01 | -.04 to .05 | .79 |
| Model 3 | .0002 | -.04 to .04 | .99 |
| Model 4 | -.01 | -.05 to .03 | .57 |
| Model 5 | -.02 | -.06 to .02 | .41 |

Multiply imputed models (100 imputations). N=3,305 individuals who had data on all three cognitive measures at age 24 in ALSPAC. 95% CI = 95% Confidence Interval. Model 1: unadjusted; Model 2: adjusted for sex, ethnicity, and BMI at age 24; Model 3: additionally adjusted for maternal education and SES; Model 4: additionally adjusted for smoking and alcohol use at age 24; Model 5: additionally adjusted for IQ at age 8. Exposure and outcome are standardised.

### 3.2. One-sample bidirectional Mendelian randomization in ALSPAC

#### 3.2.1. MR assumptions

Regression models examined whether genetic risk scores, based on the SNP instrument set, predicted the relevant exposures (circulating levels of inflammatory markers and cognitive performance) in ALSPAC. As outcome variables (CRP and IL-6) were highly skewed, they were log transformed. All instruments had *F*-statistics >10 (range:15.8 to 112.1), indicating adequate instrument strength (Table 3) (Burgess & Thompson, 2011; Staiger & Stock, 1997). Re-running the analysis removing individuals with high CRP levels (>10mg/l) did not substantially change the results, suggesting that these individuals are not driving associations.

**Table 3.** Association between genetic risk scores and exposures within ALSPAC.

| Outcome | Instrument | $F$ | $R^2$ | N |
| --- | --- | --- | --- | --- |
| **Inflammation** |  |  |  |  |
| *Primary analysis* |  |  |  |  |
| Log CRP (age 24) | Ligthart et al. (*cis*) | 26.7 | 1.2% | 2,222 |
|  | Han et al. (*cis*) | 15.8 | 0.7% | 2,222 |
| Log IL-6 (age 9) | Ahluwalia et al. (*cis*) | 87.6 | 2.1% | 4,184 |
|  | Rosa et al. (*cis*) | 76.1 | 1.8% | 4,184 |
| GlycA (age 24) | Borges et al. (genome-wide) | 65.7 | 2.7% | 2,392 |
| *Secondary analysis* |  |  |  |  |
| Log CRP (age 24) | Ligthart et al. (genome-wide) | 99.9 | 4.3% | 2,222 |
|  | Han et al. (genome-wide) | 79.9 | 3.5% | 2,222 |
| Log IL-6 (age 9) | Ahluwalia et al. (genome-wide) | 77.3 | 1.8% | 4,184 |
|  | Sarwar et al. (*cis*) | 87.9 | 2.1% | 4,184 |
|  | Swerdlow et al. (*cis*) | 95.4 | 2.2% | 4,184 |
| GlycA (age 24) | Kettunen et al. (genome-wide) | 51.8 | 2.1% | 2,392 |
| **Cognition** |  |  |  |  |
| Working memory (age 24) | Mahedy et al. | 59.7 | 2.3% | 2,534 |
| Emotion recognition (age 24) | Mahedy et al. | 112.1 | 4.1% | 2,626 |
| Response inhibition (age 24) | Mahedy et al. | 112.1 | 4.3% | 2,508 |

Separate regression models examined whether genetic risk scores were associated with potential confounders (Davies et al., 2018; Yang et al., 2022). Top 10 genetic principal components were added to all models to adjust for genetic ancestry. There was evidence that one CRP instrument (Han-genome-wide) was associated with maternal education (*p=*.00006) and alcohol use (*p*=.015); and one cognition instrument (Mahedy-emotion recognition) was associated with maternal SES (*p*=.013). This violates one of the MR assumptions and thus caution should be taken when interpreting findings using these two instruments. Evidence for other instruments was weak (Supplementary Table S13).

#### 3.2.2. Potential causal relationships between inflammatory markers and cognitive measures

We did not find strong evidence of a causal effect of genetically-proxied CRP, IL-6, sIL-6R or GlycA on standard deviation change in cognitive measures in ALSPAC at age 24 in our primary analysis using *cis* variants and larger GlycA GWAS (beta range: -0.73 [95% CI -2.47 to 1.01, *p*=.41] to 0.21 [95% CI = -1.42 to 1.84, *p*=.80]) or secondary analysis using genome-wide significant variants and smaller GWAS (beta range: -0.73 [95% CI = -2.67 to 1.21, *p*=.46] to 1.54 [95% CI = -0.34 to 3.42, *p*=.11]; N range: 1,677 to 2,193; Fig. 2 and Supplementary Table S14;). There was also not strong evidence of a causal effect of cognition on standard deviation change in inflammatory markers (*p*s ≥.19; Supplementary Table S15).

**Fig. 2.**
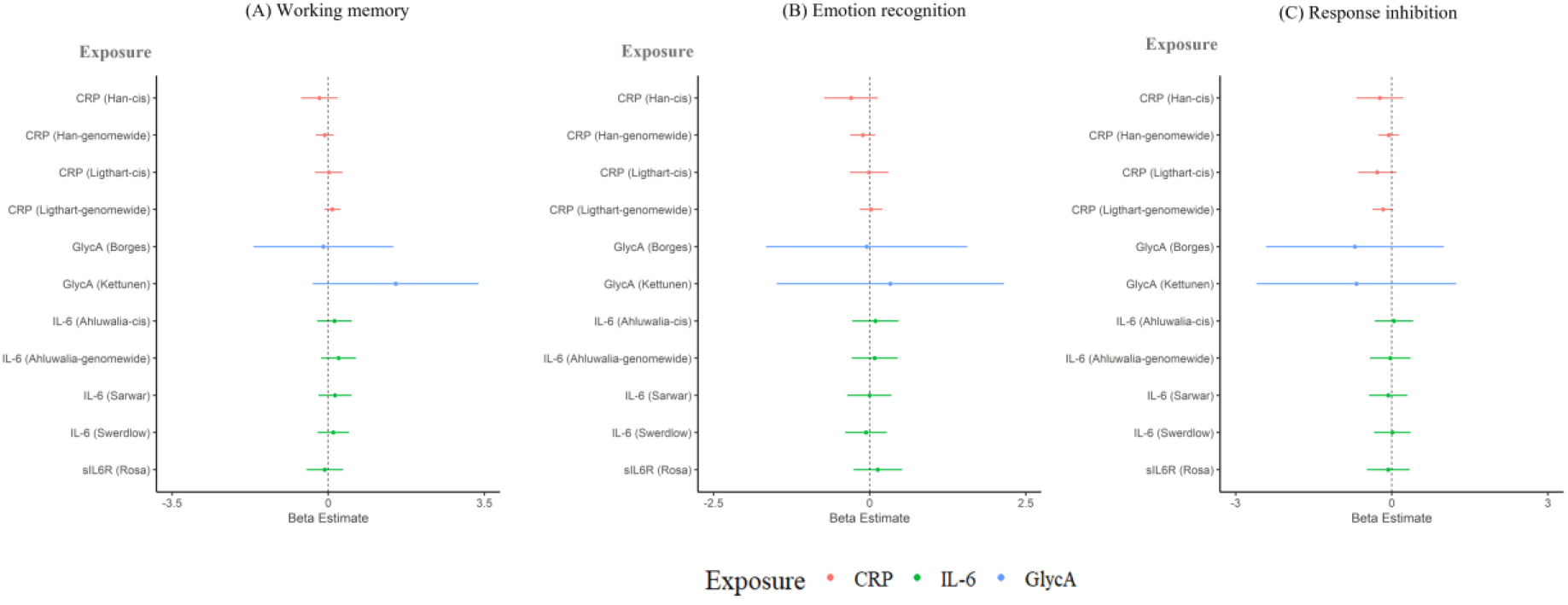
One-sample MR: effect of genetically-proxied inflammatory markers on (A) working memory, (B) emotion recognition, and (C) response inhibition at age 24 years. Outcomes are standardised. Points represent beta estimates and 95% CI. For values, see Supplementary Table S14.

### 3.3. Two-sample bidirectional Mendelian randomization

#### 3.3.1. Effect of inflammatory markers on general cognitive ability

In the primary analysis, there was not strong evidence for a causal effect of genetically-proxied inflammatory markers (CRP, IL-6, sIL6-R and GlycA) on GCA (IVW estimates range: -0.02 [95% CI = -0.05 to 0.01, *p*=.12] to 0.03 [95% CI = -0.01 to 0.07, *p*=.19]; Fig.3 and Supplementary Table S17). However, there was a pattern towards CRP increasing GCA and GlycA decreasing GCA, which was consistent amongst most sensitivity analyses. In the secondary analyses, there was some evidence for a causal effect of genetically-proxied IL-6 (Swerdlow 3 SNP instrument) on higher cognition (IVW estimate: 0.05 [95% CI = 0.02 to 0.09, *p*=.006]) and CRP on poorer cognition (IVW estimate: -0.03 [95% CI = -0.04 to -0.01, *p*=.01), although the direction of effects did not replicate across different MR methods (Fig. 4; Supplementary Table S17). Steiger filtering showed that one CRP instrument (Han – 494 genome-wide SNPs) had four invalid variants (i.e., variants had stronger associations with the outcome than the exposure). Re-running the analysis with these variants removed did not substantially change the results.

**Fig. 3.**
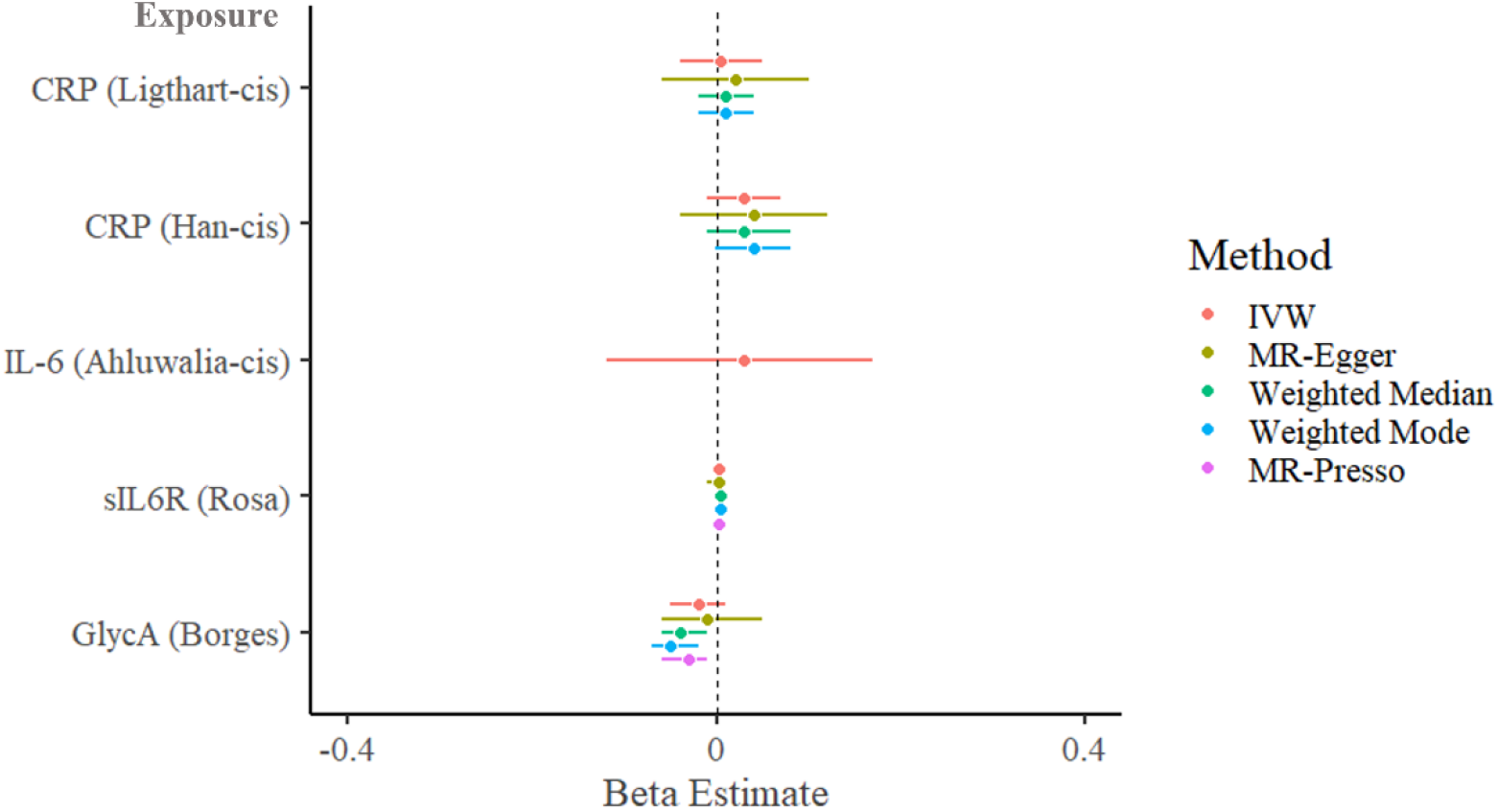
Two-sample MR (primary analysis - *cis* variants for CRP, IL-6 and sIL-6R; larger GlycA GWAS): causal effect of genetically-proxied inflammatory markers on general cognitive ability. Points represent beta estimates and 95% CI.

**Fig. 4.**
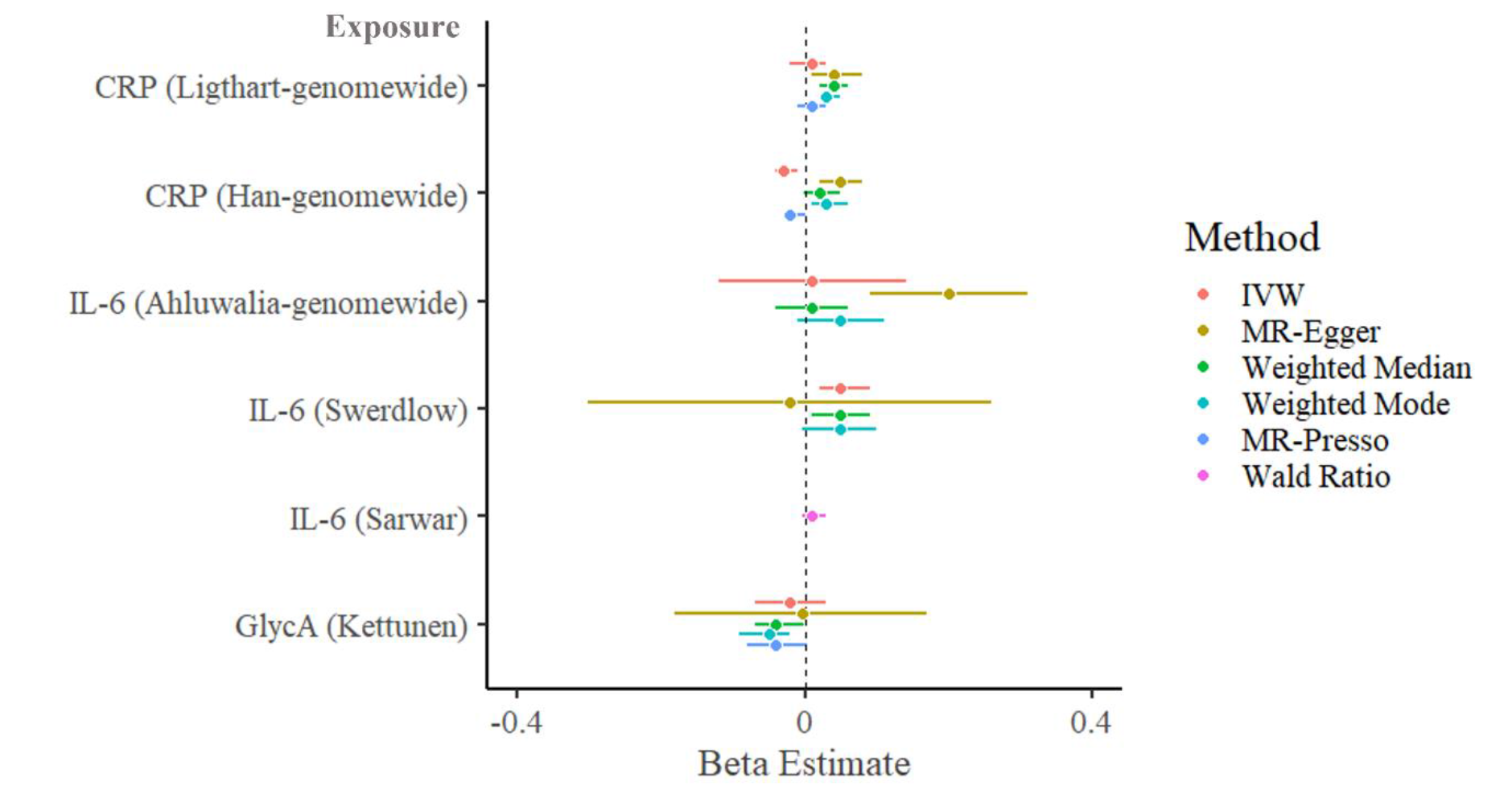
Two-sample MR (secondary analysis – genome-wide significant variants and smaller GWAS): causal effect of genetically-proxied inflammatory markers on general cognitive ability. Points represent beta estimates and 95% CI.

#### 3.3.2. Effect of general cognitive ability on inflammatory markers

In the primary analysis, there was some evidence of a causal effect of higher GCA on lower CRP (Ligthart IVW estimate: -0.11, 95% CI = -0.16 to -0.07, *p*<.0001]; Han IVW estimate: -0.02, 95% CI = -0.04 to -0.01, *p*=.005), IL-6 (Ahluwalia IVW estimate: -0.05, 95% CI = -0.09 to -0.002, *p*=.039) and GlycA (Borges IVW estimate: -0.21, 95% CI = -0.27 to - 0.16, *p*<.0001) (Fig. 5; Supplementary Table S20). Effect estimates were broadly consistent across sensitivity analyses. These findings are consistent with Steiger filtered results (Supplementary Table S22).

**Fig. 5.**
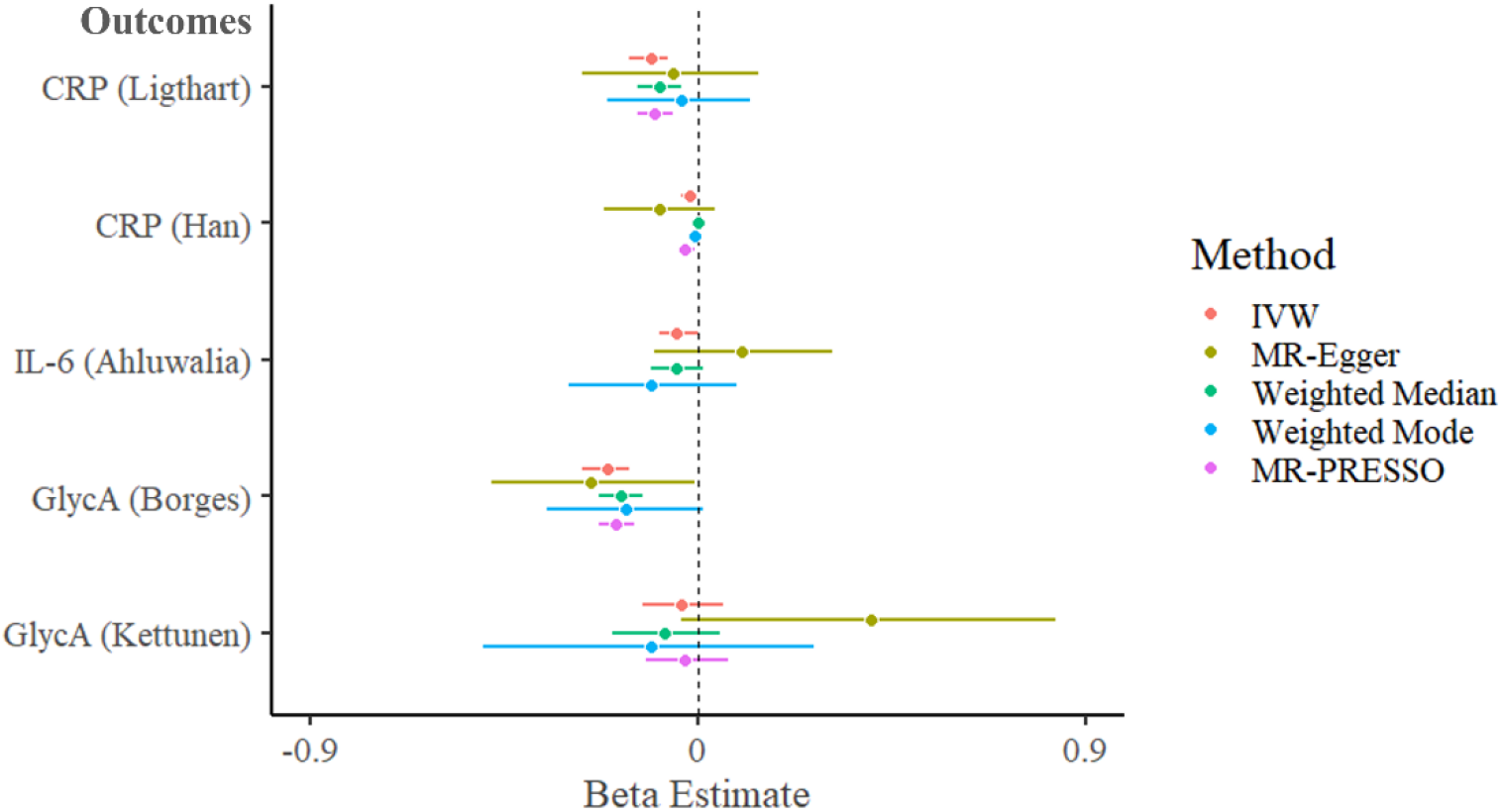
Two-sample MR: causal effect of genetically-proxied general cognitive ability on inflammatory markers. Points represents beta estimates and 95% CI.

#### 3.3.3. Assessment of heterogeneity and horizontal pleiotropy

For most instruments, there was evidence of heterogeneity based on Cochran’s *Q*-statistic (range: 0.4 to 1634). There was limited evidence of horizontal pleiotropy for inflammation (*p*s ≥ 0.42) and cognition (*p*s ≥ 0.048) instruments based on the Egger intercept, although this was not always consistent with MR-PRESSO Global Test results (Supplementary Tables S18, S21). Instruments used in secondary analyses revealed evidence of horizontal pleiotropy using the Egger intercept, highlighting the importance of using *cis* variants which may be less likely to be pleiotropic and therefore may provide more valid instruments.

## 4. Discussion

We examined associations between inflammatory markers and cognition using a large population-based cohort and complementary MR analyses. Our cross-sectional analyses show that GlycA, but not CRP, is associated with poorer working memory, emotion recognition, and response inhibition at 24 years. However, this association was fully explained by confounders, namely sex, ethnicity, BMI, maternal education, maternal SES, smoking and alcohol use, childhood IQ. In one-sample MR, there was limited evidence of a causal relationship between inflammatory markers (CRP, GlycA, IL-6, sIL-6R) and the same cognitive measures in ALSPAC, although confidence intervals were large. In two-sample MR, we did not find strong evidence of a causal effect of the same inflammatory markers on GCA. However, there was a pattern towards CRP being associated with higher GCA and GlycA being associated with poorer GCA. There was evidence that higher GCA may be causally related to lower inflammation, with the strongest evidence for GlycA.

### 4.1. Comparison with previous studies

#### 4.1.1. Inflammatory markers and specific cognitive domains

We found limited evidence of a causal relationship between inflammatory markers and cold cognitive domains (working memory and response inhibition) in early adulthood. Recent observational studies report that CRP and IL-6 (age 9) predict working memory performance one year later in ALSPAC (Kokosi et al., 2021; Shields et al., 2021). Unlike the current study, these studies focused on associations in childhood; although there are other differences which may also account for the discrepant findings (e.g., confounders included, a larger sample size). Consistent with our findings, Proitsi and colleagues (2018) did not find associations between GlycA and a related cognitive domain (short-term memory) in late midlife after adjusting for potential confounders (Proitsi et al., 2018). Moreover, experimental studies did not find consistent evidence that acute inflammatory challenges influence working memory in early adulthood (Bollen et al., 2017). As for response inhibition, to our knowledge, few observational studies have examined its association with inflammation. Experimentally, there is limited evidence that acute inflammation influences response inhibition in early adulthood, as assessed by the Stroop and Go/No-Go test (Bollen et al., 2017; Handke et al., 2020). In relation to the overarching domain of executive functioning, Mac Giollabhui and colleagues (2021) reported that CRP was associated with reduced executive functioning in 43,896 individuals aged 18-93 after adjusting for confounders (Mac Giollabhui et al., 2021). Importantly, effect sizes were small (as noted by the authors) and performance on the task used may also reflect processing speed (Kuiper et al., 2017). Taken together, there is limited evidence that peripheral inflammatory markers impact working memory and response inhibition in early adulthood.

In relation to hot cognition, we found limited evidence in support of a causal relationship between inflammatory markers and emotion recognition. To our knowledge, there are few observational studies that have examined the association between inflammation and emotion processing. Experimentally, there is some evidence that inflammatory challenges are associated with poorer social and emotional processing (Balter et al., 2018; Bollen et al., 2017). In a double-blind placebo controlled cross-over design, Balter and colleagues (2018) reported that acute inflammation induced via a Typhoid vaccination reduced accuracy on the Reading the Mind in the Eyes Test ∼6 hours post-injection (Balter et al., 2018). However, it is important to note that these studies typically involve higher acute doses of inflammation, which differ from low-grade inflammation examined here.

Collectively, there is limited evidence of a causal relationship between systemic inflammation and cognitive domains – working memory, response inhibition, emotion recognition – in early adulthood. However, we observed large confidence intervals and consistent with previous studies (Mac Giollabhui et al., 2021), effect sizes were small. Consequently, our study may be underpowered to detect these effects (Supplementary methods). MR studies and experimental studies with larger sample sizes are needed to further interrogate this question. For MR studies, there is a need for larger GWAS on specific cognitive domains.

#### 4.1.2. Inflammatory markers and general cognitive ability

We did not find strong evidence that inflammation has a causal effect on GCA across a broad age range. Nevertheless, the pattern of associations consistently showed genetically-proxied CRP to be associated with higher GCA and genetically-proxied GlycA to be associated with poorer GCA, warranting further research. A previous study including 13,000 individuals reported that GlycA was associated with poorer GCA after adjusting for cardiometabolic correlates of cognition (van der Lee et al., 2018). In addition, Conole and colleagues (2021) report that a DNA methylation signature of CRP was associated with poorer GCA in elderly individuals (aged 72) (Conole et al., 2021). Whilst these studies provide evidence that CRP and GlycA are associated with poorer GCA, they do not establish causality, as noted by the authors. Consequently, it is possible that potential confounders or reverse causality may account for these findings. A recent MR study that explored a broad range of inflammatory markers on cognitive functioning reported three cytokines (Eotaxin, Interleukin-8 and Monocyte chemotactic protein 1) were associated with higher fluid intelligence and IL-4 associated with lower fluid intelligence (little evidence for prospective memory and reaction time) (Pagoni et al., 2022). Consistent with our study, there was limited evidence to suggest a causal effect of IL-6 on fluid intelligence; other inflammatory measures (CRP and GlycA) were not examined (Pagoni et al., 2022). This suggests that although there is limited evidence of a causal effect of the inflammatory markers examined here, there is some evidence for other inflammatory markers. Taken together, few studies have examined the potential causal effect of inflammatory markers on GCA across a broad age range. Given the pattern of findings in our study, further studies investigating the role of these inflammatory markers on cognition are needed.

There was evidence that GCA may be causally related to lower inflammation, with the strongest evidence for GlycA. Arguably, this is not surprising as measures closely related to GCA (higher educational attainment and SES) are associated with lower inflammatory markers such as CRP and IL-6 (Loucks et al., 2010; Muscatell et al., 2018; Nazmi & Victora, 2007; O’Connor et al., 2009; Pollitt et al., 2007). One potential mechanism through which GCA may impact inflammation is via health-related behaviour (e.g., physical activity, smoking): individuals with higher GCA may be more likely to engage in healthier lifestyle choices (e.g., less likely to smoke) which may result in lower levels of inflammation (Davies et al., 2019). Importantly, as GCA is associated with higher SES/education, this may provide people with the means to engage in healthier lifestyle choices (e.g., via higher income) (Friedman & Herd, 2010). An alternative mechanism could involve shared biological pathways linking inflammation and cognition (Zuber et al., 2022). However, this is less likely as we only observe the effect in one direction (GCA on inflammation, not *vice versa)*. A third possible explanation is that the results are due to chance (Type I error) and do not reflect a true casual effect. Further studies are needed to explore possible mechanisms of this relationship.

### 4.2. Strengths and Limitations

A key strength of this study is the triangulation of findings using complementary analyses (cohort and MR) which provides increased confidence in our inferences drawn. We also used large population-based data, increasing statistical power and generalisability of our findings. Additionally, we examined several inflammatory markers, including a novel marker (GlycA) which may better reflect chronic inflammation. Moreover, in the cohort analyses we considered many potential confounders and in the MR analysis, we used strong instrumental variables making weak instrument bias unlikely and checked key assumptions with no primary instruments being associated with potential confounders.

A limitation is that similar to other cohorts, there is non-random attrition within ALSPAC (Wolke et al., 2009), which may lead to bias. For example, if inflammation and cognition and related to attrition (Supplementary Table S7), results may be biased towards the null. Second, many cognitive tasks (including those used here) have poor-to-moderate test-retest reliability therefore measurement error may mask potential associations by increasing confidence intervals (Hedge et al., 2018). Third, we relied on single measures of inflammatory markers which exhibit high intra-individual variability (Conole et al., 2021). However, we did include a novel inflammatory marker (GlycA) which is thought to provide a more stable measure of inflammation (Connelly et al., 2017; Ritchie et al., 2015). Fourth, MR focuses on lifetime effects of inflammation on cognition. This is informative for understanding lifetime risk of an exposure, but cannot discern effects at specific ages (i.e., potential sensitive periods). Experimental research and MR studies with genetic variants associated with inflammatory markers at specific ages (e.g., early childhood, adulthood) are required to address this temporal aspect. Fifth, we focus on a subset of cognitive domains and inflammatory markers. Future studies should examine other domains (e.g., executive functioning, reward processing) and inflammatory markers (Boyle et al., 2019; Mac Giollabhui et al., 2021). Additionally, some instruments used in the MR analyses had few SNPs, displayed evidence of heterogeneity, and/or were associated with potential confounders. Nevertheless, there was limited evidence of horizontal pleiotropy (MR-Egger intercept) or associations with potential confounders in primary analyses using *cis* variants.

### 4.3. Implications

Our study highlights the need for large-scale GWAS on specific cognitive domains. This will enable MR studies to examine causal relationships between inflammation and specific domains. Additionally, we found limited evidence that the inflammatory markers CRP, IL-6, sIL-6R and GlycA influence cognition (working memory, response inhibition, emotion recognition, GCA), suggesting that they may not be good intervention targets for poorer cognition (although further evidence is needed to determine this). Furthermore, we found a potential causal effect of GCA on inflammation and highlight the need for mechanistic studies investigating this relationship.

## 5. Conclusions

In summary, we used cross-sectional association and MR analyses to examine the association and potential causal relationship between inflammatory markers and cognition (general and domain-specific) using data from a large population-based cohort (ALSPAC) and publicly available GWAS. We did not find strong evidence for a causal effect of inflammatory markers on specific cognitive domains in young adults in the ALSPAC cohort (working memory, response inhibition, emotion recognition) or GCA. There was some evidence that GCA may be causally related to lower inflammation. There is a need for larger GWAS on specific cognitive domains, experimental studies with larger sample sizes, and studies investigating potential mechanisms of GCA on inflammation.

## Data Availability

Any researcher can apply to use ALSPAC data, including the variables under investigation in this study, see http://www.bristol.ac.uk/alspac/researchers/access/.
For availability of GWAS full summary statistics used in this study, see Supplementary Materials.

## Acknowledgements

This work is supported by the Elizabeth Blackwell Institute for Health Research, University of Bristol, and the Wellcome Trust Institutional Strategic Support Fund (204813/Z/16/Z). This research was funded in whole, or in part, by the Wellcome Trust [Grant number 204813/Z/16/Z]. For the purpose of open access, the authors have applied a CC BY public copyright licence to any Author Accepted Manuscript version arising from this submission.

This work was also supported by the Medical Research Council (MRC) Integrative Epidemiology Unit at the University of Bristol (MC_UU_00011/1, MC_UU_00011/3, MC_UU_00011/7). GMK acknowledges funding support from the Wellcome Trust (Grant No. 201486/Z/16/Z), and the UK Medical Research Council (Grant No. MC_PC_17213, Grant No. MR/S037675/1, and Grant No. MR/W014416/1). HJ is supported by the NIHR Biomedical Research Centre at University Hospitals Bristol and Weston NHS Foundation Trust and the University of Bristol. The views expressed are those of the authors and not necessarily those of the NIHR or the Department of Health and Social Care. GDS and KT work in the MRC Integrative Epidemiology Unit at the University of Bristol which is supported by the Medical Research Council (MC_UU_00011/1 and MC_UU_00011/3). CD is supported by an MRC Integrative Epidemiology Unit Fellowship (MC_UU_00011/1).

The UK Medical Research Council and Wellcome (Grant ref: 217065/Z/19/Z) and the University of Bristol provide core support for ALSPAC. This publication is the work of the authors and CS will serve as a guarantor for the contents of this paper. A comprehensive list of grants funding is available on the ALSPC website (grant-acknowledgements.pdf (bristol.ac.uk); This research was specifically funded by Wellcome Trust and MRC (Grant Ref: 076467/Z/05/Z; MR/L022206/1). GWAS data was generated by Sample Logistics and Genotyping Facilities at Wellcome Sanger Institute and LabCorp (Laboratory Corporation of America) using support from 23andMe.

We are extremely grateful to all the families who took part in this study, the midwives for their help in recruiting them, and the whole ALSPAC team, which includes interviewers, computer and laboratory technicians, clerical workers, research scientists, volunteers, managers, receptionists, and nurses.

Members of the CHARGE Inflammation Working Group are Emelia Benjamin, Daniel I. Chasman, Abbas Dehghan, Tarunveer Singh Ahluwalia, James Meigs, Russell Tracy, Behrooz Z. Alizadeh, Symen Ligthart, Josh Bis, Gudny Eiriksdottir, Nathan Pankratz, Myron Gross, Alex Rainer, Harold Snieder, James G. Wilson, Bruce M. Psaty, Josee Dupuis, Bram Prins, Urmo Vaso, Maria Stathopoulou, Lude Franke, Terho Lehtimaki, Wolfgang Koenig, Yalda Jamshidi, Sophie Siest, Ali Abbasi, Andre G. Uitterlinden, Mohammadreza Abdollahi, Renate Schnabel, Ursula M. Schick, Ilja M. Nolte, Aldi Kraja, Yi-Hsiang Hsu, Daniel S. Tylee, Alyson Zwicker, Rudolf Uher, George Davey Smith, Alanna C. Morrison, Andrew Hicks, Cornelia M. van Duijn, Cavin Ward-Caviness, Eric Boerwinkle, J. Rotter, Ken Rice, Leslie Lange, Markus Perola, Eco de Geus, Andrew P. Morris, Kari Matti Makela, David Stacey, Johan Eriksson, Tim M. Frayling, and Eline P. Slagboom.

## Ethics statement

Ethical approval for the study was obtained from the ALSPAC Ethics and Law Committee (http://www.bristol.ac.uk/alspac/researchers/research-ethics/) and the Local Research Ethics Committees. Consent for biological samples has been collected in accordance with the Human Tissue Act (2004). Informed consent for the use of data collected via questionnaires and clinics was obtained from participants following the recommendations of the ALSPAC Ethics and Law Committee at the time. ALSPAC study data were collected and managed using REDCap electronic data capture tools hosted at the University of Bristol (Harris et al., 2009). REDCap (Research Electronic Data Capture) is a secure, web-based software platform designed to support data capture for research studies.

