## Supplementary Materials for "Association between inflammation and cognition: triangulation of evidence using a population-based cohort and Mendelian randomization analyses"

### Supplementary Material

### SUPPLEMENTARY METHODS

##### *ALSPAC genotype data*

ALSPAC children were genotyped using the Illumina HumanHap550 quad chip genotyping platforms by 23andme (Genome build: Build 37). Quality controls using Plink v1.07 included excluding individuals with (1) gender mismatches, (2) minimal or excessive heterozygosity, (3) missingness (> 3%), (4) insufficient sample replication (IBD < 0.8), (5) non-European ancestry. SNPs with minor allele frequency of <1%, call rate < 95%, or evidence for violations of Hardy-Weinberg equilibrium (*p* < 5E-7) were removed. Cryptic relatedness was measured as proportion of identity by descent (IBD > 0.1). Participants who passed these quality controls were retained during subsequent phasing and imputation. This resulted in 9,115 participants and 500,527 SNPs. There were 8,237 children with genotype data available after using cryptic relatedness. Imputation of genotypes was done with Impute v2.2.2 software with the 1000 genomes reference panel, resulting in 7,191,388 SNPs after MAF > .01 and info score (indicating high imputation quality) > 0.8. For further details, please visit: alspac.github.io/omics_documentation/alspac_omics_data_catalogue.html#org48674f8

##### *ALSPAC potential confounders*

**Sex.** Sex of child was obtained from birth notification.

**Ethnicity.** Child ethnic background was defined as either white or non-white. This is a derived variable (created using responses to other questions) in ALSPAC based on two questions (C800 and C801) which asked the mother to describe the race or ethnic group of herself and her partner. Child ethnic background was defined as non-white if either the mother or partners ethnic group was reported as non-white.

**BMI at age 24.** Body mass index (BMI) at age 24 is a derived variable calculated as [weight (kg)] / [height (m)$^2$].

**Maternal education.** Mother's highest educational qualification is a derived variable in ALSPAC which includes the following categories: Degree, A-level, O-level, Vocational, CSE/none. It is important to note that here we use the original raw variable (C645). ALSPAC also created another recoded variable (C645A) which puts all mothers who left education questions blank to "CSE/none", under the assumption that mothers with no educational qualifications would leave this question blank. This recoded version was not used here, instead any questions with "not known" or that were missed were recorded as missing.

**Maternal socioeconomic status (SES).** A proxy for maternal socioeconomic position – maternal occupation – was used. Maternal occupation is a derived variables in ALSPAC which is based on the Office of Population Censuses and Surveys (OPCS) job codes. This question was asked to the mothers within ALSPAC during pregnancy and includes the following categories: professional, intermediate, skilled (non-manual), skilled (manual), partly skilled, unskilled, and armed forces. Due to a small number of individuals in the group armed forces (N < 5), these individuals were removed.

**Alcohol use at age 24**. The AUDIT-C, a shortened version of the AUDIT (1), is used to identify individuals who are hazardous drinkers or have alcohol use disorders. It includes three questions: “How often did you have a drink containing alcohol in the past year”, “How many drinks did you have on a typical day when you were drinking in the past year?” and “How often did you have six or more drinks on one occasion in the past year?”. Each question is scored 0-4. Total scores range from 0-12.

**Smoking status at age 24.** Smoking status was a derived variable based on responses to multiple questions related to smoking and score on the Fagerström Test for Nicotine Dependence (FTND) (2) (FKSM1150). The following categories were created: “never smoked a whole cigarette”, “not smoked in the last 30 days”, “not a daily smoker”, “daily smoker”. Individuals who scored 1-10 on the FTND (who must be daily smokers) were grouped into one category as “daily smokers”.

**IQ at age 8.** Total IQ score (which includes verbal and performance IQ) on the Wechsler Intelligence Scale for Children (WISC) (3) was used as a measure of IQ at age 8.

***Multiple imputation***

Participants who had data on all three cognitive outcomes at age 24 were included in the analysis (N=3,305). All participants had complete data the three cognitive outcomes and sex. For details of variables included in the imputation models, please see Table S1. For each set of imputations, 100 datasets were imputed using chained equations with the *mi impute chained* command in Stata. Auxiliary variables were identified as variables that are associated with variables being imputed (i.e., the exposures and/or potential confounders; $r \geq .09$), see Table S1 for list of Auxiliary variables. All variables were included in each model.

We ran several sensitivity analyses to check the robustness of our findings from the multiple imputation models. First, the multiple imputation produced implausible values for some predictors that were not normally distributed (e.g., minus values for CRP). Whilst the goal of multiple imputation is not to predict missing values (4), we re-ran the multiple imputation using predictive mean matching to 10-nearest neighbours to check whether only including plausible values alters our findings. This did not substantially alter our findings. Second, we checked whether re-running the multiple imputation with fewer imputed datasets (N=50) substantially affected the standard errors (i.e., uncertainty associated with missing values (5,6)). If this is the case, it may suggest that more imputed datasets are required to decrease uncertainty associated with missing values. The standard errors were similar across the two analyses. Third, we re-ran the multiply imputed models separately for each outcome: working memory (N = 3,478), emotion recognition (N = 3,613) and response inhibition (N = 3,430) to check whether this altered the findings. This did not alter our overall conclusions.

##### ***Mendelian randomization: brief description***

Mendelian randomization (MR) is a method used to assess causality (7,8). This method uses genetic variants (typically Single Nucleotide Polymorphisms; SNPs) strongly associated with environmental exposures of interest as proxies for the exposure (8). MR is less susceptible to the limitations of conventional epidemiological approaches (reverse causation and confounding), and if certain assumptions are met, allows causal inferences between the exposure and outcome to be drawn (8).

MR can be conducted in a one or two sample setting (9). One-sample MR involves using individual level data. In many one-sample MR studies, SNPs identified from GWAS conducted on the exposure are used to create a genetic risk score which is used to indicate lifetime risk of the exposure (10). The causal effect of the exposure on the outcome is then often assessed using two-stage least squares regression (11). Two-sample MR often uses summary-level data from publicly available GWAS. Here, SNPs are treated like individual studies (e.g., randomised controlled trials) which are then meta-analysed. There are advantages and disadvantages of both MR approaches, see (7) for more details. It is important to note the three key assumptions of MR: (a) instruments are associated with the exposure, (b) instruments are not associated with potential confounders and (c) instruments are associated with the outcome only via the exposure (10). If key assumptions are not met, this reduces confidence in inferences drawn from MR analyses. In two-sample MR, there is the additional assumption that GWAS for the exposure and outcome come from similar but not overlapping participants (9).

##### ***Additional details of GWAS used to create instruments for MR***

**Ligthart et al. (2018)** Circulating CRP was natural log transformed. Individuals were excluded from all analyses if they had an auto-immune disease, were taking immune-modulating agents (if information was available), or they had CRP ≥ 4 SD from the mean.

**Han et al. (2020)** Circulating CRP was rank-based inverse-normal transformed. Average values of serum CRP were calculated for individuals that underwent two assessments.

**Ahluwalia et al. (2021)** Circulating IL-6 was natural log-transformed. Only population-based samples or healthy controls from case-control studies were included in the final analyses.

**Borges et al. (2020)** Details not available.

**Kettunen et al. (2016)** For details on individual criteria applied in studies used in the Kettunen GWAS, see Kettunen et al. (2016) supplementary materials.

**Rosa et al. (2019)** The Rosa instrument was based on the Sun et al. (2018) GWAS on sIL-6R. The cohorts in this GWAS included participants who were generally in good health. Blood donation criteria excluded individuals with a history of major diseases (such as myocardial infarction, stroke, cancer, HIV, and hepatitis B or C) and individuals who have had recent illness or infection. For details on blood sample collections, see (17). Quality controls included exclusions for sex mismatches, low call rates, duplicate sample, extreme heterozygosity, and non-European descent.

**Swerdlow et al. (2012)** Circulating IL-6 was natural log transformed.

**Sarwar et al. (2012)** Circulating IL-6 was natural log transformed.

**Mahedy et al. (2021)** In all three cognitive GWAS (working memory, emotion recognition, response inhibition), no transformations were applied to the outcomes. In the emotion recognition and response inhibition GWAS, no exclusions were applied. In the working memory GWAS, individuals who responded to < 50% trials or had a negative score (*d*-prime) were excluded.

**Lam et al. (2021)** MTAG of two GWAS:

> **Davies et al. (2018)** A general cognitive ability score was derived from two consortia (COGENT and CHARGE) and UK Biobank. For each cohort in CHARGE and COGENT, the general cognitive function component was constructed from several cognitive tasks (required a minimum of three different domains) using principal component analysis. In UK Biobank, scores on the verbal-numerical reasoning test (13-item multiple-choice questions) that assesses 'fluid' cognitive ability was used. Details on all cognitive phenotypes from all cohorts is reported in Davies et al. (2018) supplementary Note 1. Exclusion criteria included clinical stroke (including self-reported stroke) or prevalent dementia.
>
> **Savage et al. (2018)** A general cognitive ability score was derived from each cohort (except the High IQ/Health and Retirement Study where a logistic regression was run predicting whether participants were drawn from a population of very high intelligence). Cohorts had either a single sum score, mean score, or factor score from a battery of cognitive tests (for example, IQ score, fluid intelligence test and cognitive tasks such as digit span/processing speed). For more details on cognitive tests used, and exclusion criteria applied in each cohort, see Savage et al. (2018).

***Creating weighted genetic risk scores***

Weighted genetic risk scores were created for inflammatory (CRP, IL-6, GlycA, sIL-6R) and cognitive (working memory, emotion recognition, response inhibition) phenotypes for each ALSPAC participant in Plink v1.90 (24). Specifically, risk alleles were weighted by the effect size (beta) reported in the GWAS and then summed to provide a single risk score. Unrelated individuals were kept, and withdrawals of consent were removed. For SNPs not available in ALSPAC, proxies were identified that had: $r^2 > 0.8$ (using *LDproxy_batch* function in EUR population in R), rsID available, SNP available in full summary statistics and ALSPAC. Quality checks involved (1) checking there were no mismatches in SNP alleles between base data and ALSPAC (no mismatches were detected) and (2) checking for palindromic SNPs (SNPs with alleles A/T or C/G). Palindromic SNPs have the same allele pairs on both the forward and backward strand, and therefore if the base or outcome GWAS does not specify which strand the analysis was done on, there is the possibility that they could be reporting from different strands resulting in an error in the MR results. In total, there were 88 distinct palindromic SNPs. As there were no mismatches in allele pairs between the base data and ALSPAC, it is unlikely that there are strand differences. Nevertheless, a sensitivity analysis was conducted removing these SNPs to check whether this influenced the results. Re-running the analysis with these SNPs removed did not substantially influence the results.

***Statistical power for one-sample MR***

We conducted a *post-hoc* power calculation using mRnd (25) (shiny.cnsgenomics.com/mRnd/) to check the statistical power of the one-sample MR. The following parameters were used based on the data obtained in this study: power (0.08), alpha (0.05), $\beta_{yx}$ (approximate regression coefficient from one-sample MR assuming this is the true effect size; 0.1); $\beta$OLS (approximate regression coefficient from observational analysis; 0.05), $\sigma2(x)$ and $\sigma2(y)$ (variance of exposure and outcome based on per SD change; 1). For $R^2xz$ (proportion of variance explained for the association between allele score and exposure variable), we set various thresholds given the variability across instruments (conservative = 0.02; liberal = 0.03; very liberal = 0.04). Based on this, the required sample size to detect the expected effect size would be conservative (N = 39,295), liberal (N = 26,197) and very liberal (N = 19,648).

*Two-sample MR methods*

**Inverse Variance Weighted (IVW) method**. This method is often used in meta-analyses where individual studies are weighted by the inverse of their variance (i.e., their precision) and combined to estimate an average effect (26,27). In MR, instead of individual studies, individual SNP effects (Wald ratios) are combined. Wald ratios are calculated by dividing the SNP outcome association by the SNP exposure association. This method forces the intercept through zero assuming no horizontal pleiotropy (i.e., it assumes SNPs are associated with the outcome *only* via the exposure). Therefore, the IVW method will provide a consistent estimate if all SNPs are valid instrumental variables (26,27).

**MR-Egger method.** This method is similar to the IVW method except that it does not force the intercept through zero. Consequently, this method provides an estimate in the presence of invalid SNPs (SNPs that affect the outcome through pathways other than the exposure) (28). The slope provides a causal effect estimate and the intercept can be used to indicate the degree of horizontal pleiotropy.

**Weighted-median method.** This method uses the median of the ratio estimates and will provide a consistent estimate if at least 50% of the weights come from valid SNPs (29).

**Weighted-mode method.** This method will provide a consistent estimate if the most common causal effect estimates come from valid SNPs (even if the majority of SNPs are not valid) (30).

**MR-PRESSO method.** This method consists of the following three tests: (a) MR-PRESSO global test which can be used to detect horizontal pleiotropy, (b) MR-PRESSO outlier test which removes outliers from IVW estimates, (c) MR-PRESSO distortion test which tests whether there is a large distortion in the causal estimates once outlier have been removed (31). In this study, we also used the MR-PRESSO outlier test to examine whether the causal effect estimates are robust to the removal of outliers.

### SUPPLEMENTARY TABLES

***Table S1. Variables included in the multiple imputation models (N=3,305)***

| Variable | N with missing data (% missing) | Regression model used to impute missing data |
|---|---|---|
| **Outcome variables** | | |
| Working memory (age 24, z-score) | 0 (0%) | N/A |
| Emotion recognition (age 24, z-score) | 0 (0%) | N/A |
| Response inhibition (age 24, z-score) | 0 (0%) | N/A |
| **Exposure variables** | | |
| CRP (age 24, z-score) | 807 (24.4%) | Linear |
| GlycA (age 24, z-score) | 590 (17.9%) | Linear |
| **Potential confounders** | | |
| Sex | 0 (0%) | N/A |
| Ethnicity | 344 (10.4%) | Logistic |
| BMI (age 24) | 34 (1.0%) | Linear |
| Maternal education | 380 (11.5%) | Ordinal Logistic |
| Maternal SES | 658 (19.9%) | Ordinal Logistic |
| Alcohol use (age 24) | 56 (1.7%) | Linear |
| Smoking status (age 24) | 29 (0.9%) | Ordinal Logistic |
| IQ (age 8) | 585 (17.7%) | Linear |
| **Auxiliary variables** | | |
| CRP (age 9) | 1,360 (41.2%) | Linear |
| CRP (age 15) | 1,583 (47.9%) | Linear |
| CRP (age 17) | 1,536 (46.5%) | Linear |
| GlycA (age 7) | 1,339 (40.5%) | Linear |
| GlycA (age 15) | 1,630 (49.3%) | Linear |
| GlycA (age17) | 1,589 (48.1%) | Linear |
| IL-6 (age 9) | 1,363 (41.2%) | Linear |
| Working memory (age 10) | 682 (20.6%) | Linear |
| Alcohol use (AUDIT – age 17) | 1,040 (31.5%) | Linear |
| Maternal financial difficulties (pregnancy) | 366 (11.1%) | Linear |
| Maternal age (delivery) | 221 (6.7%) | Linear |
| BMI (age 7) | 480 (14.5%) | Linear |
| BMI (age 13) | 653 (19.8%) | Linear |
| BMI (age 15) | 768 (23.2%) | Linear |
| BMI (age 17) | 698 (21.1%) | Linear |
| Maternal depression (18 weeks gestation) | 474 (14.3%) | Linear |
| Monocyte levels (age 24) | 643 (19.5%) | Linear |
| Lymphocyte levels (age 24) | 643 (19.5%) | Linear |
| Paternal SES | 504 (15.3%) | Ordinal Logistic |

***Table S2. Characteristics of ALSPAC participants.***

| Phenotype | ALSPAC variable name | Descriptive statistics *Mean (SD), min max* | N |
|---|---|---|---|
| | | **Exposures** | |
| CRP (age 24) | CRP_F24 | *M* = 2.28 (6.52)<br>Min = .1<br>Max = 224.72 | 3,015 |
| | CRP_F24 (excludes values ≥ 10 mg/l) | *M* = 1.55 (1.82)<br>Min = .1<br>Max = 9.8 | 2,901 |
| GlycA (age 24) | Gp_F24 | *M* = 1.23 (0.17)<br>Min = 0.84<br>Max = 2.25 | 3,258 |
| | Gp_F24 (excludes CRP_F24 ≥ 10) | *M* = 1.22 (0.16)<br>Min = 0.84<br>Max = 2.19 | 3,144 |
| | | **Outcomes** | |
| Emotion recognition (age 24) | FKEP1070 | *M* = 66.36 (7.89)<br>Min = 25<br>Max = 88 | 3,613 |
| Working memory (age 24) | Derived variable | *M* = 2.76 (0.80)<br>Min = 0<br>Max = 3.78 | 3,478 |
| Response inhibition (age 24) | FKEP3060 | *M* = 258.72 (53.11)<br>Min = 67<br>Max = 508 | 3,430 |
| | | **Potential Confounders** | |
| BMI (age 24) | FKMS1040 | *M* = 24.92 (5.08)<br>Min = 13.68<br>Max = 63.74 | 3,974 |
| Alcohol use (age 24) | FKAL1500 | *M* = 5.16 (2.51)<br>Min = 0<br>Max = 12 | 3,928 |
| IQ (age 8) | f8ws112 | *M* = 103.97 (16.54)<br>Min = 45<br>Max = 151 | 7,346 |
| Sex | kz021 | 0. Male = 7,690<br>1. Female = 7,348 | 15,038 |
| Ethnicity | c804 | 0. White = 11,523<br>1. Non-white = 613 | 12,136 |
| Maternal education | c645 | 0. Degree = 1,608<br>1. A level = 2,793 | 11,703 |

| | | | |
|---|---|---|---|
| | | 2. O level = 4,323<br>3. Vocational = 1,229<br>4. CSE/none = 1,750 | |
| Maternal SEP | c755 | 0. Professional = 595<br>1. Intermediate = 3,180<br>2. Skilled (non-manual) = 4,322<br>3. Skilled (manual) = 790<br>4. Partly skilled = 997<br>5. Unskilled = 222 | 10,106 |
| Smoking status (age 24) | FKSM1150 | 0. Never smoked a whole cigarette = 1,435<br>1. Not smoked in last 30 days = 1,390<br>2. Not a daily smoker = 642<br>3. Daily smoker = 486 | 3,953 |

N = excludes missing data; maternal education = mothers highest education qualification; maternal SEP = maternal occupation using Office of Population Censuses and Surveys (OPCS) job codes as a proxy for socioeconomic position; CSE = certificate of secondary education.

***Table S3. Cross-sectional association between CRP and cognitive outcomes at age 24 in ALSPAC, unadjusted and adjusted for potential confounders (complete cases).***

| Outcome (models) | *b* | 95% CI | *p*-value | N |
|---|---|---|---|---|
| **Working Memory** | | | | |
| Model 1 | -.03 | -.06, .009 | .14 | 2,624 |
| Model 2 | -.02 | -.06, .02 | .34 | 2,327 |
| Model 3 | -.03 | -.08, .03 | .33 | 2,020 |
| Model 4 | -.02 | -.08, .03 | .39 | 1,976 |
| Model 5 | -.01 | -.06, .04 | .69 | 1,700 |
| **Emotion Recognition** | | | | |
| Model 1 | -.01 | -.05, .02 | .49 | 2,718 |
| Model 2 | -.01 | -.05, .03 | .54 | 2,407 |
| Model 3 | .002 | -.05, .05 | .94 | 2,089 |
| Model 4 | .003 | -.05, .06 | .91 | 2,042 |
| Model 5 | .01 | -.04, .06 | .67 | 1,758 |
| **Response Inhibition** | | | | |
| Model 1 | .02 | -.02, .06 | .29 | 2,582 |
| Model 2 | .004 | -.03, .04 | .83 | 2,291 |
| Model 3 | .01 | -.04, .07 | .65 | 1,988 |
| Model 4 | .003 | -.05, .06 | .91 | 1,945 |
| Model 5 | -.001 | -.05, 05 | .98 | 1,686 |

95% CI = 95% Confidence Interval. Model 1: unadjusted; Model 2: adjusted for sex, ethnicity, and BMI at age 24; Model 3: additionally adjusted for maternal education and socioeconomic position; Model 4: additionally adjusted for smoking and alcohol use at age 24; Model 5: additionally adjusted for IQ at age 8. Exposure and outcomes are standardised.

***Table S4. Cross-sectional association between GlycA and cognitive outcomes at age 24 in ALSPAC, unadjusted and adjusted for potential confounders (complete cases).***

| Outcome (models) | *b* | 95% CI | *p*-value | N |
|---|---|---|---|---|
| **Working Memory** | | | | |
| Model 1 | -.08 | -.11, -.04 | <.001 | 2,849 |
| Model 2 | -.06 | -.10, -.02 | .004 | 2,523 |
| Model 3 | -.04 | -.08, .005 | .083 | 2,190 |
| Model 4 | -.03 | -.08, .02 | .19 | 2,142 |
| Model 5 | -.02 | -.06, .03 | .53 | 1,839 |
| **Emotion Recognition** | | | | |
| Model 1 | -.05 | -.09, -.01 | .007 | 2,949 |
| Model 2 | -.02 | -.06, .03 | .45 | 2,610 |
| Model 3 | .007 | -.04, .05 | .77 | 2,266 |
| Model 4 | .003 | -.04, .05 | .88 | 2,215 |
| Model 5 | .009 | -.04, .06 | .72 | 1,902 |
| **Response Inhibition** | | | | |
| Model 1 | .05 | .008, .08 | .016 | 2,806 |
| Model 2 | .02 | -.02, .06 | .34 | 2,487 |
| Model 3 | .009 | -.04, .05 | .71 | 2,158 |
| Model 4 | -.002 | -.05, .05 | .94 | 2,111 |
| Model 5 | -.02 | -.07, .03 | .50 | 1,825 |

95% CI = 95% Confidence Interval. Model 1: unadjusted; Model 2: adjusted for sex, ethnicity, and BMI at age 24; Model 3: additionally adjusted for maternal education and socioeconomic position; Model 4: additionally adjusted for smoking and alcohol use at age 24; Model 5: additionally adjusted for IQ at age 8. Exposure and outcomes are standardised.

***Table S5. Cross-sectional association between CRP and cognitive outcomes at age 24 in ALSPAC, unadjusted and adjusted for potential confounders, excluding individuals with CRP > 10mg/l (complete cases).***

| Outcome (model) | *b* | 95% CI | *p*-value | N |
|---|---|---|---|---|
| **Working Memory** | | | | |
| Model 1 | -.05 | -.09, -.01 | .008 | 2,527 |
| Model 2 | -.02 | -.06, .03 | .48 | 2,247 |
| Model 3 | -.01 | -.06, .04 | .69 | 1,956 |
| Model 4 | -.01 | -.06, .04 | .69 | 1,915 |
| Model 5 | .008 | -.04, .06 | .74 | 1,644 |
| **Emotion Recognition** | | | | |
| Model 1 | -.02 | -.06, .02 | .36 | 2,617 |
| Model 2 | -.001 | -.04, .04 | .97 | 2,323 |
| Model 3 | .005 | -.04, .05 | .84 | 2,022 |
| Model 4 | .002 | -.04, .05 | .94 | 1,978 |
| Model 5 | .002 | -.05, .05 | .95 | 1,699 |
| **Response Inhibition** | | | | |
| Model 1 | .03 | -.01, .07 | .19 | 2,490 |
| Model 2 | -.01 | -.06, .03 | .63 | 2,215 |
| Model 3 | -.02 | -.06, .03 | .53 | 1,926 |
| Model 4 | -.02 | -.07, .03 | .48 | 1,885 |
| Model 5 | -.03 | -.08, .03 | .33 | 1,630 |

95% CI = 95% Confidence Interval. Model 1: unadjusted; Model 2: adjusted for sex, ethnicity, and BMI at age 24; Model 3: additionally adjusted for maternal education and socioeconomic position; Model 4: additionally adjusted for smoking and alcohol use at age 24; Model 5: additionally adjusted for IQ at age 8. Exposure and outcomes are standardised.

***Table S6. Cross-sectional association between GlycA and cognitive outcomes at age 24 in ALSPAC, unadjusted and adjusted for potential confounders, excluding individuals with CRP > 10mg/l (complete cases).***

| Outcome (model) | *b* | 95% CI | *p*-value | N |
|---|---|---|---|---|
| **Working Memory** | | | | |
| Model 1 | -.06 | -.10, -.03 | .001 | 2,752 |
| Model 2 | -.05 | -.09, -.01 | .024 | 2,443 |
| Model 3 | -.03 | -.07, .02 | .22 | 2,126 |
| Model 4 | -.02 | -.07, .03 | .42 | 2,081 |
| Model 5 | -.005 | -.05, .04 | .85 | 1,783 |
| **Emotion Recognition** | | | | |
| Model 1 | -.04 | -.08, -.01 | .020 | 2,848 |
| Model 2 | -.01 | -.05, .03 | .67 | 2,526 |
| Model 3 | .01 | -.04, .05 | .73 | 2,199 |
| Model 4 | .003 | -.04, .05 | .90 | 2,151 |
| Model 5 | .01 | -.04, .06 | .73 | 1,843 |
| **Response Inhibition** | | | | |
| Model 1 | .03 | -.01, .06 | .19 | 2,714 |
| Model 2 | .01 | -.04, .05 | .75 | 2.411 |
| Model 3 | -.01 | -.05, .04 | .77 | 2,096 |
| Model 4 | -.02 | -.06, .03 | .51 | 2,051 |
| Model 5 | -.03 | -.08, .02 | .28 | 1,769 |

95% CI = 95% Confidence Interval. Model 1: unadjusted; Model 2: adjusted for sex, ethnicity, and BMI at age 24; Model 3: additionally adjusted for maternal education and socioeconomic position; Model 4: additionally adjusted for smoking and alcohol use at age 24; Model 5: additionally adjusted for IQ at age 8. Exposure and outcome are standardised.

***Table S7. Logistic regression to predict missingness in cognitive data at age 24.***

| Predictor | *b* | 95% CI | *p* | *N* |
| --- | --- | --- | --- | --- |
| **IQ – age 8** | -0.03 | -0.03 to -0.03 | <0.001 | 7,346 |
| **Sex (ref. male)** | -0.70 | -0.78 to -0.62 | <0.001 | 15,038 |
| **Ethnicity (ref. white)** | 0.34 | 0.13 to 0.55 | 0.001 | 12,136 |
| **BMI – age 24** | 0.03 | 0.02 to 0.05 | <0.001 | 3,974 |
| **Maternal education** | | | | 11,703 |
| Degree | [reference] | [reference] | [reference] | |
| A level | 0.39 | 0.27 to 0.52 | <0.001 | |
| O level | 0.82 | 0.69 to 0.94 | <0.001 | |
| Vocational | 1.19 | 1.01 to 1.37 | <0.001 | |
| CSE | 1.65 | 1.48 to 1.83 | <0.001 | |
| **Maternal socioeconomic position** | | | | 10,106 |
| Professional | [reference] | [reference] | [reference] | |
| Intermediate | 0.44 | 0.26 to 0.62 | <0.001 | |
| Skilled (non-manual) | 0.84 | 0.66 to 1.01 | <0.001 | |
| Skilled (manual) | 1.19 | 0.95 to 1.44 | <0.001 | |
| Partly skilled | 1.30 | 1.06 to 1.53 | <0.001 | |
| Unskilled | 1.50 | 1.09 to 1.92 | <0.001 | |
| **Smoking – age 24** | | | | 3,953 |
| Never smoked | [reference] | [reference] | [reference] | |
| Not smoked last 30 days | -0.07 | -0.27 to 0.14 | 0.52 | |
| Not daily smoker | 0.27 | 0.03 to 0.52 | 0.027 | |
| Daily smoker | 0.63 | 0.38 to 0.88 | <0.001 | |
| **Alcohol – age 24** | -0.05 | -0.08 to -0.01 | 0.005 | 3,928 |
| **CRP – age 24** | 0.01 | -0.01 to 0.02 | 0.35 | 3,015 |
| **GlycA – age 24** | 0.83 | 0.31 to 1.34 | 0.002 | 3,258 |
| **CRP – age 9** | 0.0004 | -0.02 to 0.02 | 0.97 | 5,080 |
| **CRP – age 15** | 0.013 | -0.01 to 0.03 | 0.16 | 3,488 |
| **CRP – age 17** | 0.01 | -0.01 to 0.02 | 0.27 | 3,285 |

| | | | | |
|---|---|---|---|---|
| **GlycA – age 7** | -0.40 | -0.79 to -0.003 | 0.048 | 5,518 |
| **GlycA – age 15** | 0.09 | -0.43 to 0.62 | 0.72 | 3,363 |
| **GlycA – age 17** | 0.70 | 0.18 to 1.21 | 0.008 | 3,173 |
| **IL-6 – age 9** | -0.02 | -0.05 to 0.02 | 0.37 | 5,070 |

For each participant in ALSPAC, the outcome was coded as either 0 (not missing – individual has data on all three cognitive tasks at age 24; N = 3,305) or 1 (missing – individual does not have data on all three cognitive tasks at age 24).

***Table S8. Details of GWAS used to create instruments for one and two sample MR.***

| **Phenotype** | **GWAS/ Instrument** | **Population** | **Cohort/ studies(*s*)** | **Covariates** | **Ages** | **N** | **Includes ALSPAC** (approximate % sample if applicable) | **Ref** |
|---|---|---|---|---|---|---|---|---|
| **CRP** | Ligthart et al. (2018) | European ancestry | 88 studies | Adjusted for age, sex, population structure, accounting for relatedness, if relevant. | Cohorts range from *M* age of 9.9 to 86.6 years | 204,402 | Yes (HapMap not 1KG GWAS)<br>ALSPAC (N = 4,099) in total sample (N = 204,402) = **2%.** | (12) |
| | Han et al. (2020) | European ancestry | UK Biobank | Adjusted for sex, age and first ten principal components. | *M* = 56.8 years (*SD* = 8.01) | 418,642 | No. | (13) |
| **IL-6** | Ahluwalia et al. (2021) | European ancestry | 26 cohorts | Adjusted for age, sex, population substructure (through study-specific principal components) and/or study-specific site, when necessary. | Cohorts range from *M* age of 9.9 to 86.6 years | 52,654 | Yes. ALSPAC (N = 4,129) in discovery sample (N = 52,654) = **7.8%** | (14) |
| | Swerdlow et al. (2012) Instrument | European ancestry | Whitehall II study | Identified SNPs *a priori,* then tested the association between SNPs and log IL-6 in Whitehall II. | *M* = 49.2 years (*SD* = 6.0) | Up to 4,479 per SNP | No. | (18) |

| | | | | | | | | |
|---|---|---|---|---|---|---|---|---|
| | Sarwar et al. (2012) Instrument | European ancestry (≥ 90%) | 16 studies | Unknown | Unknown | 27,185 | Unknown | (19) |
| **GlycA** | Borges et al. (2020) | European | UK Biobank | Unknown | Unknown | 115,078 | No | N/A |
| | Kettunen et al. (2016) | European | 14 cohorts | All metabolites were adjusted for age, sex, time from last meal, if applicable, and ten first principal components from genomic data and the resulting residuals were transformed to normal distribution by inverse rank-based normal transformation. | Cohorts range from *M* age 23.9 to 61.3 years. | 19,270 | No. | (15) |
| **sIL6R** | Rosa et al. (2019) Instrument from Sun et al., (2018) GWAS on sIL6R. | European ancestry | INTERVAL study (UK) | Adjusted for sex, age, duration between blood draw and processing, first 3 ancestry principal components. | Cohorts *M* age is 44 years (*SD* = 14) | 3,301 | No. | (16,32) |
| **Working memory** | Mahedy et al. (2021) | European | ALSPAC | Adjusted for age, sex, and first 10 genetic principal components. | 24 years | 2,471 | Yes. | (20) |
| **Emotion recognition** | Mahedy et al. (2021) | European | ALSPAC | Adjusted for age, sex, and first 10 genetic principal components | 24 years | 2,560 | Yes. | (20) |

| | | | | | | | | |
|---|---|---|---|---|---|---|---|---|
| **Response Inhibition** | Mahedy et al. (2021) | European | ALSPAC | Adjusted for age, sex, and first 10 genetic principal components | 24 years | 2,446 | Yes. | (20) |
| **General Cognitive Ability** | *Lam et al. (2021) | European ancestry | Combined two cognitive GWAS: Savage et al. (2018) (14 cohorts) and Davies et al. (2018) (57 cohorts), with ~ 89% sample overlap. | Davies et al. (2018) adjusted for age, sex, and population stratification were included in the model for each cohort. Cohort-specific covariates (site or familial relationships) were also fitted as required.<br><br>Savage et al. (2018) adjusted for age, sex, ancestry principal components. | Davies et al. (2018) cohorts ages range from 16 to 102 years.<br><br>Savage et al. (2018) cohorts ages range from 5 to 98 years. | 373,617 | No. | (21) |

*M* = mean; CRP = C-reactive protein; IL-6 = Interleukin-6; GlycA = Glycoprotein acetyls; sIL-6R = soluble interleukin-6 receptor; ALSPAC = Avon Longitudinal Study of Parents and Children; GWAS = genome-wide association studies; unknown = information not reported in paper (to authors knowledge). * = not all cohorts from the two cognitive GWAS were included due to problems with data access.

***Table S9. Variance explained by SNPs in original GWAS paper for comparison.***

| Original paper | Variance explained by SNPs |
|---|---|
| Ligthart et al. (2018) | Same sample: lead variants at distinct loci explained up to 7.0% variance in CRP levels. Additional detail: 52-SNPs (48: HapMap, 4: 1KG GWAS): $R^2 = 0.065$, *F*-statistic = 273. |
| Han et al. (2020) | Same sample: 526 SNPs explained 13% variance in CRP levels. |
| Ahluwalia et al. (2021) | Independent sample (NESDA): three GWAS index SNPs explained ~ 1.06% variance in IL-6 in NESDA cohort (rs4537545, rs660895, rs6734238). |
| Kettunen et al. (2016) | Same sample: 74 variants (associated with one or more metabolic traits) explained 2.41% variance in glycoprotein acetyls. |
| Rosa et al. (2019) | Same sample: 34 *cis* SNPs ($r^2 < 0.1$, F-statistic > 15) located within 250kb IL6R. *F*-statistic estimates ($beta^2/SE^2$) for individual SNPs predicting sIL6R ranged from 15.73 to 504.90 |

Instruments reported in original papers may not contain the same SNPs that were used as instruments in this paper due to different criterion applied; NESDA = Netherlands Study of Depression and Anxiety; SNP = Single Nucleotide Polymorphism; same sample = variance explained by SNPs in same sample used to conduct GWAS; independent sample = variance explained by SNPs in an independent sample to that used to conduct GWAS.

***Table S10. Source of GWAS full summary statistics and instruments***

| GWAS Full Summary Statistics/Instruments | Source | Link (if available online) or author contact details |
|---|---|---|
| Ligthart et al. (2018) | IEU Open GWAS Project (https://gwas.mrcieu.ac.uk/) | gwas.mrcieu.ac.uk/datasets/ieu-b-35/ |
| Han et al. (2020) | Requested from authors | Corresponding author: Xikun Han |
| Ahluwalia et al. (2021) | Requested from authors | Corresponding authors: Tarunveer Ahluwalia Behrooz Alizadeh |
| Swerdlow et al. (2012)* | Taken from Nils Kappelmann OSF | OSF: osf.io/apme9/ |
| Sarwar et al. (2012) | Taken from Nils Kappelmann OSF | OSF: osf.io/apme9/ |
| Borges et al. (2020) | IEU Open GWAS Project | gwas.mrcieu.ac.uk/datasets/met-d-GlycA/ |
| Kettunen et al. (2016) | IEU Open GWAS Project | gwas.mrcieu.ac.uk/datasets/met-c-863/ |
| Rosa et al. (2019) | Available in paper supplementary | www.nature.com/articles/s41525-019-0097-4#Sec30 |
| Mahedy et al. (2021) (Working memory) | University of Bristol Open Repository | research-information.bris.ac.uk/en/datasets/genome-wide-association-of-working-memory |
| Mahedy et al. (2021) (Emotion recognition) | University of Bristol Open Repository | research-information.bris.ac.uk/en/datasets/genome-wide-association-study-of-emotion-recognition |
| Mahedy et al. (2021) (Response inhibition) | University of Bristol Open Repository | research-information.bris.ac.uk/en/datasets/genome-wide-association-study-of-response-inhibition |
| Lam et al. (2021) | Requested from authors | Corresponding author: Todd Lencz |

GWAS taken from IEU Open GWAS Project were converted from Variant Call Format (VCF) to text files using BCF tools (33); OSF = Open Science Framework; * = error in effect alleles reported in paper, corrected version used instead.

***Table S11. Number of SNPs available from each GWAS after criterion applied.***

| GWAS | SNPs met *p*-value criteria | Independent SNPs | Quality check | Genome-wide | *Cis* |
|---|---|---|---|---|---|
| Ligthart et al. (2018) | 3,950 | 78 | 78 | 78 | 6 |
| Han et al. (2020) | 60,177 | 552 | 552 | 552 | 20 |
| Ahluwalia et al. (2021) | 94 | 3 | 3 | 3 | 2 |
| Borges et al. (2020) | 15,328 | 88 | 87 | 87 | N/A |
| Kettunen et al. (2016) | 315 | 10 | 10 | 10 | N/A |
| Mahedy et al. (2021) (Working memory) | 6* | 3 | 3 | 3 | N/A |
| Mahedy et al. (2021) (Emotion recognition) | 15* | 6 | 6 | 6 | N/A |
| Mahedy et al. (2021) (Response inhibition) | 16* | 6 | 6 | 6 | N/A |
| Lam et al. (2021) | 16,696 | 250 | 250 | 250 | N/A |

Some instruments were not extracted from GWAS full summary statistics (i.e., already available instruments) and so they are not included here: Rosa et al. 2019, Swerdlow et al. 2012, Sarwar et al. 2012; SNPs met *p*-value criteria = SNPs with $p < 5 \times 10^{-8}$ (*except for cognitive GWAS where a less stringent criteria was applied: $p < 5 \times 10^{-6}$); Independent SNPs = SNPs met clumping criteria ($r^2 = 0.01$, kb = 1000); Quality check = SNPs with minor allele frequency > 0.01; *Cis* = SNPs located +/- 1-mB of protein coding gene; Genome-wide = SNPs that met statistical criteria. Location of protein coding gene is based on Genome Reference Consortium Human (GRCh) 37 for CRP (chr1:159,682,079-159,684,379; consistent with SNP base pair (BP) positions in CRP GWAS). As the IL-6 GWAS (Ahluwalia et al., 2021) SNP BP positions were based on GRCh36, the BP position for these SNPs were extracted from GRCh38 along with the corresponding location of the IL6R (chr1:154,405,193-154,469,450).

***Table S12. One-sample MR in ALSPAC: Number of SNPs with proxies included.***

| Exposure Instrument | SNPs that met criteria | SNPs available in ALSPAC | SNPs missing in ALSPAC | SNPs that met criteria (+ proxies) | SNPs (+ proxies) available in ALSPAC | Final N SNPs in genetic risk score |
|---|---|---|---|---|---|---|
| Ligthart et al. (*cis*) | 6 | 6 | 0 | 6 | 6 | 6 |
| Ligthart et al. (genome-wide) | 78 | 76 | 2 | 77 | 76 | 76 |
| Han et al. (*cis*) | 20 | 18 | 2 | 18 | 18 | 18 |
| Han et al. (genome-wide) | 552 | 509 | 43 | 529 | 520 | 520 |
| Ahluwalia et al. (*cis*) | 2 | 2 | 0 | 2 | 2 | 2 |
| Ahluwalia et al. (genome-wide) | 3 | 3 | 0 | 3 | 3 | 3 |
| Borges et al. | 87 | 78 | 9 | 84 | 82 | 82 |
| Kettunen et al. | 10 | 10 | 0 | 10 | 10 | 10 |
| Rosa et al. | 34 | 34 | 0 | 34 | 34 | 34 |
| Swerdlow et al. | 3 | 3 | 0 | 3 | 3 | 3 |
| Sarwar et al. | 1 | 1 | 0 | 1 | 1 | 1 |
| Mahedy et al. (Working memory) | 3 | 3 | 0 | 3 | 3 | 3 |
| Mahedy et al. (Emotion recognition) | 6 | 6 | 0 | 6 | 6 | 6 |
| Mahedy et al. (Response inhibition) | 6 | 6 | 0 | 6 | 6 | 6 |

***Table S13. One-sample MR in ALSPAC: association between genetic risk scores and potential confounders in linear regression models.***

| Exposure Instrument | Potential Confounders | N |
|---|---|---|
| Ligthart et al. (*cis*) | Sex (p = .47) | 8,114 |
| | Ethnicity (p = .89) | 7,172 |
| | BMI at age 24 (p = .34) | 2,849 |
| | Maternal education (p = .79) | 6,951 |
| | Maternal socioeconomic position (p = .63) | 6,158 |
| | Smoking at age 24 (p = .54) | 2,845 |
| | Alcohol use at age 24 (p = .99) | 2,824 |
| Ligthart et al. (genome-wide) | Sex (p = .98) | 8,114 |
| | Ethnicity (p = .80) | 7,172 |
| | BMI at age 24 (p = .49) | 2,849 |
| | Maternal education (p = .075) | 6,951 |
| | Maternal socioeconomic position (p = .13) | 6,158 |
| | Smoking at age 24 (p = .22) | 2,845 |
| | Alcohol use at age 24 (p = .13) | 2,824 |
| Han et al. (*cis*) | Sex (p = .82) | 8,114 |
| | Ethnicity (p = .58) | 7,172 |
| | BMI at age 24 (p = .37) | 2,849 |
| | Maternal education (p = .91) | 6,951 |
| | Maternal socioeconomic position (p = .22) | 6,158 |
| | Smoking at age 24 (p = .78) | 2,845 |
| | Alcohol use at age 24 (p = .57) | 2,824 |
| Han et al. (genome-wide) | Sex (p = .98) | 8,114 |
| | Ethnicity (p = .91) | 7,172 |
| | BMI at age 24 (p = .064) | 2,849 |
| | Maternal education (p = .00006) | 6,951 |
| | Maternal socioeconomic position (p = .28) | 6,158 |
| | Smoking at age 24 (p = .51) | 2,845 |
| | Alcohol use at age 24 (p = .015) | 2,824 |
| Ahluwalia et al. (*cis*) | Sex (p = .52) | 8,114 |
| | Ethnicity (p = .86) | 7,172 |
| | BMI at age 24 (p = .83) | 2,849 |
| | Maternal education (p = .77) | 6,951 |
| | Maternal socioeconomic position (p = .57) | 6,158 |
| | Smoking at age 24 (p = .47) | 2,845 |
| | Alcohol use at age 24 (p = .79) | 2,824 |
| Ahluwalia et al. (genome-wide) | Sex (p = .35) | 8,114 |
| | Ethnicity (p = .55) | 7,172 |
| | BMI at age 24 (p = .99) | 2,849 |
| | Maternal education (p = .49) | 6,951 |
| | Maternal socioeconomic position (p = .34) | 6,158 |
| | Smoking at age 24 (p = .55) | 2,845 |
| | Alcohol use at age 24 (p = .69) | 2,824 |

| | | |
|---|---|---|
| Borges et al. | Sex (p = .91) | 8,114 |
| | Ethnicity (p = .96) | 7,172 |
| | BMI at age 24 (p = .37) | 2,849 |
| | Maternal education (p = .17) | 6,951 |
| | Maternal socioeconomic position (p = .52) | 6,158 |
| | Smoking at age 24 (p = .28) | 2,845 |
| | Alcohol use at age 24 (p = .074) | 2,824 |
| Kettunen et al. | Sex (p = .96) | 8,114 |
| | Ethnicity (p = .19) | 7,172 |
| | BMI at age 24 (p = .69) | 2,849 |
| | Maternal education (p = .62) | 6,951 |
| | Maternal socioeconomic position (p = .79) | 6,158 |
| | Smoking at age 24 (p = .30) | 2,845 |
| | Alcohol use at age 24 (p = .76) | 2,824 |
| Rosa et al. | Sex (p = .90) | 8,114 |
| | Ethnicity (p = .90) | 7,172 |
| | BMI at age 24 (p = .49) | 2,849 |
| | Maternal education (p =.51) | 6,951 |
| | Maternal socioeconomic position (p = .86) | 6,158 |
| | Smoking at age 24 (p = .62) | 2,845 |
| | Alcohol use at age 24 (p = .52) | 2,824 |
| Swerdlow et al. | Sex (p = .35) | 8,114 |
| | Ethnicity (p = .25) | 7,172 |
| | BMI at age 24 (p = .81) | 2,849 |
| | Maternal education (p = .36) | 6,951 |
| | Maternal socioeconomic position (p = .66) | 6,158 |
| | Smoking at age 24 (p = .69) | 2,845 |
| | Alcohol use at age 24 (p = .98) | 2,824 |
| Sarwar et al. | Sex (p = .64) | 8,114 |
| | Ethnicity (p = .62) | 7,172 |
| | BMI at age 24 (p = .84) | 2,849 |
| | Maternal education (p = .52) | 6,951 |
| | Maternal socioeconomic position (p = .65) | 6,158 |
| | Smoking at age 24 (p = .37) | 2,845 |
| | Alcohol use at age 24 (p = .78) | 2,824 |
| Mahedy et al. (Working memory) | Sex (p = .45) | 8,114 |
| | Ethnicity (p = .31) | 7,172 |
| | BMI at age 24 (p = .20) | 2,849 |
| | Maternal education (p = .11) | 6,951 |
| | Maternal socioeconomic position (p = .60) | 6,158 |
| | Smoking at age 24 (p = .88) | 2,845 |
| | Alcohol use at age 24 (p = .30) | 2,824 |

| | | |
|---|---|---|
| Mahedy et al. (Emotion recognition) | Sex (p = .98) | 8,114 |
| | Ethnicity (p = .70) | 7,172 |
| | BMI at age 24 (p = .061) | 2,849 |
| | Maternal education (p = .55) | 6,951 |
| | Maternal socioeconomic position (p = .013) | 6,158 |
| | Smoking at age 24 (p = .62) | 2,845 |
| | Alcohol use at age 24 (p = .84) | 2,824 |
| Mahedy et al. (Response inhibition) | Sex (p = .96) | 8,114 |
| | Ethnicity (p = .18) | 7,172 |
| | BMI at age 24 (p = .30) | 2,849 |
| | Maternal education (p = .99) | 6,951 |
| | Maternal socioeconomic position (p = .30) | 6,158 |
| | Smoking at age 24 (p = .30) | 2,845 |
| | Alcohol use at age 24 (p = .40) | 2,824 |

All models include top 10 genetic principal components to adjust for population stratification.

***Table S14. One-sample MR in ALSPAC: effect of inflammatory markers on standard deviation change in cognition.***

| Outcome | Exposure | Exposure GRS | Estimate | *SE* | *p* | *N* |
|---|---|---|---|---|---|---|
| | | **Primary analysis** | | | | |
| Working memory | Log CRP | Ligthart et al. (*cis*) | 0.003 | 0.16 | 0.99 | 1963 |
| | | Han et al. (*cis*) | -0.22 | 0.21 | 0.29 | 1963 |
| | Log IL-6 | Ahluwalia et al. (*cis*) | 0.19 | 0.20 | 0.35 | 1694 |
| | | Rosa et al. | -0.05 | 0.22 | 0.82 | 1694 |
| | GlycA | Borges et al. | -0.22 | 0.81 | 0.79 | 2122 |
| Emotion recognition | Log CRP | Ligthart et al. (*cis*) | -0.02 | 0.16 | 0.92 | 2029 |
| | | Han et al. (*cis*) | -0.31 | 0.22 | 0.15 | 2029 |
| | Log IL-6 | Ahluwalia et al. (*cis*) | 0.08 | 0.19 | 0.69 | 1751 |
| | | Rosa et al. | 0.12 | 0.20 | 0.55 | 1751 |
| | GlycA | Borges et al. | 0.21 | 0.83 | 0.80 | 2193 |
| Response inhibition | Log CRP | Ligthart et al. (*cis*) | -0.27 | 0.19 | 0.16 | 1939 |
| | | Han et al. (*cis*) | -0.21 | 0.23 | 0.36 | 1939 |
| | Log IL-6 | Ahluwalia et al. (*cis*) | 0.01 | 0.20 | 0.94 | 1677 |
| | | Rosa et al. | -0.10 | 0.22 | 0.63 | 1677 |
| | GlycA | Borges et al. | -0.73 | 0.89 | 0.41 | 2098 |
| | | **Secondary analysis** | | | | |
| Working memory | Log CRP | Ligthart et al. (genome-wide) | 0.10 | 0.10 | 0.31 | 1963 |
| | | Han et al. (genome-wide) | -0.06 | 0.10 | 0.55 | 1963 |
| | Log IL-6 | Ahluwalia et al. (genome-wide) | 0.28 | 0.21 | 0.18 | 1694 |
| | | Swerdlow et al. | 0.15 | 0.19 | 0.42 | 1694 |
| | | Sarwar et al. | 0.20 | 0.20 | 0.33 | 1694 |
| | GlycA | Kettunen et al. | 1.54 | 0.96 | 0.11 | 2122 |
| Emotion recognition | Log CRP | Ligthart et al. (genome-wide) | -0.001 | 0.09 | 0.99 | 2029 |
| | | Han et al. (genome-wide) | -0.14 | 0.10 | 0.16 | 2029 |
| | Log IL-6 | Ahluwalia et al. (genome-wide) | 0.06 | 0.20 | 0.78 | 1751 |
| | | Swerdlow et al. | -0.08 | 0.18 | 0.66 | 1751 |
| | | Sarwar et al. | -0.03 | 0.19 | 0.87 | 1751 |
| | GlycA | Kettunen et al. | 0.37 | 0.94 | 0.69 | 2193 |
| Response inhibition | Log CRP | Ligthart et al. (genome-wide) | -0.18 | 0.10 | 0.073 | 1939 |
| | | Han et al. (genome-wide) | -0.08 | 0.10 | 0.43 | 1939 |
| | Log IL-6 | Ahluwalia et al. (genome-wide) | -0.05 | 0.20 | 0.80 | 1677 |
| | | Swerdlow et al. | -0.02 | 0.19 | 0.94 | 1677 |
| | | Sarwar et al. | -0.10 | 0.20 | 0.63 | 1677 |
| | GlycA | Kettunen et al. | -0.73 | 0.99 | 0.46 | 2098 |

Two-stage least squares regression; GRS = genetic risk score; Log CRP = natural log transformed CRP at age 24; Log IL-6 = natural log transformed IL-6 at age 9; GlycA = GlycA at age 24. Outcome measures are standardised (i.e., estimates reflect per standard deviation change in outcome to enable comparison across cognitive domains). Models include top 10 genetic principal components.

***Table S15. One-sample MR in ALSPAC: effect of cognitive functioning on standard deviation change in inflammatory markers.***

| **Outcome** | **Exposure** | **Exposure GRS** | **Estimate** | ***SE*** | ***p*** | ***N*** |
|---|---|---|---|---|---|---|
| Log CRP | Working memory (age 24) | Mahedy et al. (Working memory) | -0.03 | 0.19 | 0.88 | 1963 |
| | Emotion recognition (age 24) | Mahedy et al. (Emotion recognition) | -0.01 | 0.02 | 0.54 | 2029 |
| | Response inhibition (age 24) | Mahedy et al. (Response inhibition) | 0.0003 | 0.002 | 0.87 | 1939 |
| Log IL-6 | Working memory (age 24) | Mahedy et al. (Working memory) | 0.30 | 0.25 | 0.22 | 1694 |
| | Emotion recognition (age 24) | Mahedy et al. (Emotion recognition) | -0.02 | 0.02 | 0.33 | 1751 |
| | Response inhibition (age 24) | Mahedy et al. (Response inhibition) | 0.003 | 0.002 | 0.19 | 1677 |
| GlycA | Working memory (age 24) | Mahedy et al. (Working memory) | 0.14 | 0.20 | 0.47 | 2122 |
| | Emotion recognition (age 24) | Mahedy et al. (Emotion recognition) | 0.001 | 0.02 | 0.95 | 2193 |
| | Response inhibition (age 24) | Mahedy et al. (Response inhibition) | -0.0005 | 0.002 | 0.81 | 2098 |

Two stage least squares regression; GRS = genetic risk score; Log CRP = natural log transformed CRP at age 24; Log IL-6 = natural log transformed IL-6 at age 9; GlycA = GlycA at age 24. All outcome measures are standardised (i.e., estimates reflect standard deviation change in outcome to enable comparisons across outcomes). Models include top 10 genetic principal components.

***Table S16. Two-sample MR (inflammatory markers on general cognitive ability): Number of SNPs with proxies included.***

| Exposure Instrument | SNPs that met statistical criteria | SNPs missing in Outcome GWAS (Lam et al.) | Proxies for missing SNPs | Excluded SNPs | Final N SNPs used (proxies included) |
|---|---|---|---|---|---|
| Ligthart et al. (*cis*) | 6 | 0 | N/A | 0 | 6 |
| Ligthart et al. (genome-wide) | 78 | 4 | 3 | 0 | 77 |
| Han et al. (*cis*) | 20 | 9 | 2 | 0 | 13 |
| Han et al. (genome-wide) | 552 | 108 | 50 | 0 | 494 |
| Ahluwalia et al. (*cis*) | 2 | 0 | N/A | 0 | 2 |
| Ahluwalia et al. (genome-wide) | 3 | 0 | N/A | 0 | 3 |
| Borges et al. | 87 | 14 | 9 | 0 | 82 |
| Kettunen et al. | 10 | 1 | 1 | 0 | 10 |
| Rosa et al. | 34 | 7 | N/A | 5 | 22 |
| Swerdlow et al. | 3 | 0 | N/A | 0 | 3 |
| Sarwar et al. | 1 | 0 | N/A | 0 | 1 |

N/A = not applicable. For Rosa et al., it was not possible to obtain proxy SNPs because GWAS full summary statistics were not used; Excluded SNPs = palindromic SNPs with intermediate effect allele frequencies excluded as it is not possible to infer strand.

***Table S17. Two-sample MR: effect of inflammatory markers on general cognitive ability.***

| Phenotype | Genetic Instrument | MR method | *b* | 95% CI | *p* |
|---|---|---|---|---|---|
| **Primary analysis** | | | | | |
| CRP | Ligthart (*cis*) 6 SNPs | IVW | 0.005 | -0.04 to 0.05 | 0.82 |
| | | MR-Egger | 0.02 | -0.06 to 0.10 | 0.63 |
| | | Weighted Median | 0.01 | -0.02 to 0.04 | 0.44 |
| | | Weighted Mode | 0.01 | -0.02 to 0.04 | 0.53 |
| | | MR-PRESSO | N/A | N/A | N/A |
| | Han (*cis*) 13 SNPs | IVW | 0.03 | -0.01 to 0.07 | 0.19 |
| | | MR-Egger | 0.04 | -0.04 to 0.12 | 0.36 |
| | | Weighted Median | 0.03 | -0.01 to 0.08 | 0.10 |
| | | Weighted Mode | 0.04 | -0.0005 to 0.08 | 0.077 |
| | | MR-PRESSO | N/A | N/A | N/A |
| IL-6 | Ahluwalia (cis) 2 SNPs | IVW | 0.03 | -0.12 to 0.17 | 0.72 |
| sIL6R | Rosa 22 SNPs | IVW | 0.003 | -0.002 to 0.01 | 0.22 |
| | | MR-Egger | 0.002 | -0.01 to 0.01 | 0.66 |
| | | Weighted Median | 0.004 | -0.0004 to 0.01 | 0.078 |
| | | Weighted Mode | 0.004 | -0.0001 to 0.01 | 0.069 |
| | | MR-PRESSO | 0.003 | -0.002 to 0.01 | 0.24 |
| GlycA | Borges 82 SNPs | IVW | -0.02 | -0.05 to 0.01 | 0.12 |
| | | MR-Egger | -0.01 | -0.06 to 0.05 | 0.84 |
| | | Weighted Median | -0.04 | -0.06 to -0.01 | 0.008 |
| | | Weighted Mode | -0.05 | -0.07 to -0.02 | 0.001 |
| | | MR-PRESSO | -0.03 | -0.06 to -0.01 | 0.01 |
| **Secondary analysis** | | | | | |
| CRP | Ligthart (genome-wide) 77 SNPs | IVW | 0.01 | -0.02 to 0.03 | 0.57 |
| | | MR-Egger | 0.04 | 0.01 to 0.08 | 0.026 |
| | | Weighted Median | 0.04 | 0.02 to 0.06 | 0.0003 |
| | | Weighted Mode | 0.03 | 0.02 to 0.05 | 0.0005 |
| | | MR-PRESSO | 0.01 | -0.01 to 0.03 | 0.55 |
| | Han (genome-wide) 494 SNPs | IVW | -0.03 | -0.04 to -0.01 | 0.01 |
| | | MR-Egger | 0.05 | 0.02 to 0.08 | 0.002 |
| | | Weighted Median | 0.02 | -0.0004 to 0.05 | 0.054 |
| | | Weighted Mode | 0.03 | 0.01 to 0.06 | 0.008 |
| | | MR-PRESSO | -0.02 | -0.03 to 0.0001 | 0.053 |
| IL-6 | Ahluwalia (genome-wide) 3 SNPs | IVW | 0.01 | -0.12 to 0.14 | 0.91 |
| | | MR-Egger | 0.20 | 0.09 to 0.31 | 0.18 |
| | | Weighted Median | 0.01 | -0.04 to 0.06 | 0.63 |
| | | Weighted Mode | 0.05 | -0.01 to 0.11 | 0.23 |
| | Swerdlow 3 SNPs | IVW | 0.05 | 0.02 to 0.09 | 0.006 |
| | | MR-Egger | -0.02 | -0.30 to 0.26 | 0.91 |
| | | Weighted Median | 0.05 | 0.01 to 0.09 | 0.027 |
| | | Weighted Mode | 0.05 | -0.003 to 0.10 | 0.20 |
| | Sarwar 1 SNP | Wald Ratio | 0.01 | -0.003 to 0.03 | 0.11 |
| GlycA | Kettunen 10 SNPs | IVW | -0.02 | -0.07 to 0.03 | 0.51 |
| | | MR-Egger | -0.003 | -0.18 to 0.17 | 0.97 |

| | | | | |
|---|---|---|---|---|
| | Weighted Median | -0.04 | -0.07 to -0.002 | 0.037 |
| | Weighted Mode | -0.05 | -0.09 to -0.02 | 0.013 |
| | MR-PRESSO | -0.04 | -0.08 to 0.004 | 0.13 |

***Table S18. Two-sample MR (inflammatory markers on general cognitive ability): test of heterogeneity and pleiotropy.***

| Exposure | Exposure Instrument | Method | Estimate | *p* |
|---|---|---|---|---|
| **Primary** | | | | |
| CRP | Ligthart (*cis*) | Q statistic (IVW) | 14.13 | 0.01 |
| | | Egger intercept | -0.002 | 0.65 |
| | | MR-PRESSO global test | 20.95 | 0.23 |
| | Han (*cis*) | Q statistic (IVW) | 21.70 | 0.04 |
| | | Egger intercept | -0.001 | 0.75 |
| | | MR-PRESSO global test | 26.51 | 0.08 |
| IL-6 | Ahluwalia (*cis*) | Q statistic (IVW) | 8.10 | 0.004 |
| | | Egger intercept | NA | NA |
| | | MR-PRESSO global test | NA | NA |
| | Rosa | Q statistic (IVW) | 52.13 | 0.0002 |
| | | Egger intercept | 0.0005 | 0.87 |
| | | MR-PRESSO global test | 54.82 | 0.001 |
| GlycA | Borges | Q statistic (IVW) | 356.01 | <0.0001 |
| | | Egger intercept | -0.001 | 0.42 |
| | | MR-PRESSO global test | 366.65 | <0.0001 |
| **Secondary** | | | | |
| CRP | Ligthart (genome-wide) | Q statistic (IVW) | 301.07 | <0.0001 |
| | | Egger intercept | -0.002 | 0.015 |
| | | MR-PRESSO global test | 310.91 | <0.0001 |
| | Han (genome-wide) | Q statistic (IVW) | 1576.31 | <0.0001 |
| | | Egger intercept | -0.002 | <0.0001 |
| | | MR-PRESSO global test | 1585.37 | <0.0001 |
| IL-6 | Ahluwalia (genome-wide) | Q statistic (IVW) | 14.05 | 0.0009 |
| | | Egger intercept | -0.01 | 0.17 |
| | | MR-PRESSO global test | NA | NA |
| | Swerdlow | Q statistic (IVW) | 0.37 | 0.83 |
| | | Egger intercept | 0.006 | 0.70 |
| | | MR-PRESSO global test | NA | NA |
| GlycA | Kettunen | Q statistic (IVW) | 51.15 | <0.0001 |
| | | Egger intercept | -0.002 | 0.88 |
| | | MR-PRESSO global test | 68.36 | <0.0001 |

***Table S19. Two-sample MR (general cognitive ability on inflammatory markers): Number of SNPs with proxies included.***

| Exposure Instrument (general cognitive ability) | Outcome GWAS (inflammation) | SNPs missing in outcome GWAS | Proxies for missing SNPs | Excluded SNPs | Final N SNPs used (proxies included) |
|---|---|---|---|---|---|
| Lam et al. (250 SNPs) | Ligthart et al. | 121 | 90 | 0 | 219 |
| | Han et al. | 1 | 0 | 0 | 249 |
| | Ahluwalia et al. | 113 | 85 | 0 | 222 |
| | Borges et al. | 0 | 0 | 0 | 250 |
| | Kettunen et al. | 2 | 2 | 1 | 249 |

Excluded SNPs = palindromic SNPs with intermediate effect allele frequencies (minor allele frequency > 0.42) excluded as it is not possible to infer strand.

***Table S20. Two-sample MR: effect of general cognitive ability on inflammatory markers.***

| **Phenotype** | **Outcome GWAS** | **MR method** | ***b*** | **95% CI** | ***p*** |
|---|---|---|---|---|---|
| CRP | Ligthart (219 SNPs) | IVW | -0.11 | -0.16 to -0.07 | <0.0001 |
| | | MR-Egger | -0.06 | -0.27 to 0.14 | 0.55 |
| | | Weighted Median | -0.09 | -0.14 to -0.04 | 0.0006 |
| | | Weighted Mode | -0.04 | -0.21 to 0.12 | 0.60 |
| | | MR-PRESSO | -0.10 | -0.14 to -0.06 | <0.0001 |
| | Han (249 SNPs) | IVW | -0.02 | -0.04 to -0.01 | 0.005 |
| | | MR-Egger | -0.09 | -0.22 to 0.04 | 0.16 |
| | | Weighted Median | -0.001 | -0.01 to 0.01 | 0.77 |
| | | Weighted Mode | -0.01 | -0.02 to 0.01 | 0.34 |
| | | MR-PRESSO | -0.03 | -0.04 to -0.01 | 0.0004 |
| IL-6 | Ahluwalia (222 SNPs) | IVW | -0.05 | -0.09 to -0.002 | 0.039 |
| | | MR-Egger | 0.10 | -0.10 to 0.31 | 0.33 |
| | | Weighted Median | -0.05 | -0.11 to 0.01 | 0.098 |
| | | Weighted Mode | -0.11 | -0.30 to 0.09 | 0.28 |
| | | MR-PRESSO | N/A | N/A | N/A |
| GlycA | Borges (250 SNPs) | IVW | -0.21 | -0.27 to -0.16 | <0.0001 |
| | | MR-Egger | -0.25 | -0.48 to -0.01 | 0.040 |
| | | Weighted Median | -0.18 | -0.23 to -0.13 | <0.0001 |
| | | Weighted Mode | -0.17 | -0.35 to 0.01 | 0.061 |
| | | MR-PRESSO | -0.19 | -0.23 to -0.15 | <0.0001 |
| | Kettunen (249 SNPs) | IVW | -0.04 | -0.13 to 0.06 | 0.45 |
| | | MR-Egger | 0.40 | -0.04 to 0.83 | 0.078 |
| | | Weighted Median | -0.08 | -0.20 to 0.05 | 0.22 |
| | | Weighted Mode | -0.11 | -0.50 to 0.27 | 0.57 |
| | | MR-PRESSO | -0.03 | -0.12 to 0.07 | 0.58 |

***Table S21. Two-sample MR (general cognitive ability on inflammatory markers): test of heterogeneity and pleiotropy.***

| Exposure Instrument | Outcome GWAS | Method | Estimate | *p* |
|---|---|---|---|---|
| Lam et al. (2021) | Ligthart et al. | Q statistic (IVW) | 437.23 | <0.0001 |
| | | Egger intercept | -0.0009 | 0.63 |
| | | MR-PRESSO global test | 442.01 | <0.0001 |
| | Han et al. | Q statistic (IVW) | 1633.52 | <0.0001 |
| | | Egger intercept | 0.001 | 0.27 |
| | | MR-PRESSO global test | 1740.46 | <0.0001 |
| | Ahluwalia et al. | Q statistic (IVW) | 285.07 | 0.002 |
| | | Egger intercept | -0.003 | 0.15 |
| | | MR-PRESSO global test | 287.84 | 0.003 |
| | Borges et al. | Q statistic (IVW) | 728.38 | <0.0001 |
| | | Egger intercept | 0.0006 | 0.78 |
| | | MR-PRESSO global test | 735.04 | <0.0001 |
| | Kettunen et al. | Q statistic (IVW) | 339.46 | 0.0001 |
| | | Egger intercept | -0.008 | 0.048 |
| | | MR-PRESSO global test | 342.22 | 0.0002 |

***Table S22. Two-sample MR: effect of general cognitive ability on inflammatory markers following Steiger filtering.***

| Phenotype | Outcome GWAS | MR method | *b* | 95% CI | *p* |
|---|---|---|---|---|---|
| CRP | Ligthart (218 SNPs) | IVW | -0.11 | -0.15 to -0.06 | <0.0001 |
| | | MR-Egger | -0.09 | -0.29 to 0.11 | 0.38 |
| | | Weighted Median | -0.09 | -0.13 to -0.04 | 0.0004 |
| | | Weighted Mode | -0.05 | -0.21 to 0.12 | 0.60 |
| | Han (246 SNPs) | IVW | -0.02 | -0.03 to -0.01 | 0.005 |
| | | MR-Egger | -0.12 | -0.23 to -0.0001 | 0.051 |
| | | Weighted Median | -0.001 | -0.01 to 0.01 | 0.77 |
| | | Weighted Mode | -0.001 | -0.01 to 0.01 | 0.84 |
| IL-6 | Ahluwalia (215 SNPs) | IVW | -0.03 | -0.07 to 0.01 | 0.14 |
| | | MR-Egger | 0.09 | -0.11 to 0.28 | 0.38 |
| | | Weighted Median | -0.04 | -0.10 to 0.02 | 0.15 |
| | | Weighted Mode | -0.11 | -0.32 to 0.10 | 0.29 |
| GlycA | Borges (239 SNPs) | IVW | -0.17 | -0.21 to -0.13 | <0.0001 |
| | | MR-Egger | -0.20 | -0.37 to -0.04 | 0.014 |
| | | Weighted Median | -0.16 | -0.21 to -0.11 | <0.0001 |
| | | Weighted Mode | -0.17 | -0.37 to 0.03 | 0.10 |
| | Kettunen (191 SNPs) | IVW | 0.006 | -0.09 to 0.10 | 0.91 |
| | | MR-Egger | 0.19 | -0.23 to 0.62 | 0.37 |
| | | Weighted Median | -0.05 | -0.18 to 0.08 | 0.46 |
| | | Weighted Mode | -0.14 | -0.55 to 0.27 | 0.51 |

***Table S23. Deviations from pre-registration with justifications***

| Deviation | Justification |
|---|---|
| **Cross-sectional analysis** | |
| Regression models weighted by missingness were not applied. | We did not run this analysis as we do not believe this would add to the current findings. |
| **One-sample MR** | |
| One instrument for CRP (CCGC) replaced with more recent GWAS for CRP. | During instrument acquisition and prior to analysis, we decided not to include the CCGC GWAS due to the availability of larger more recent GWAS (13). To increase statistical power, this GWAS was used instead. |
| **Two-sample MR** | |
| Sensitivity analysis (Generalised summary-based MR, MR-Raps) not applied. Instead, we applied Steiger filtering. | Based on the primary results, we did not run this analysis as we did not feel this would add to the current findings. |

***Table S24. Code used for analysis in this paper.***

| Study | Description | Language/ Operating system | File Name |
|---|---|---|---|
| **Cross-sectional analysis (ALSPAC)** | Cleans variables, runs multiple regression models, and creates dataset for multiple imputation. | *Stata (16)* | "InfCog_CrossSection_ALSPAC_OA.do" |
| | Runs multiple imputation, and imputed regression models. | *Stata (16)* | "InfCog_CrossSection_MI_ALSPAC_OA.do" |
| **One-sample MR (ALSPAC)** | Extracts inflammation SNPs | *R (4.1.1)* | "InfCog_1SMR_InflammationInstruments_OA.R" |
| | Extracts cognition SNPs | *R (4.1.1)* | "InfCog_1SMR_CognitionInstruments_OA.R" |
| | Checks for allele mismatches and palindromic SNPs | *R (4.1.1)* | "InfCog_1SMR_QC_AmbigSNPs_Strand_OA.R" |
| | Bash scripts create genetic risk scores using Plink | *Linux* | "InfCog_1SMR_PRS _Script1_OA.sh"<br>"InfCog_1SMR_PRS _Script2_OA.sh"<br>"InfCog_1SMR_PRS_ Script3_OA.sh" |
| | One-sample MR analysis | *R (4.1.1)* | "InfCog_1SMR_Analysis_ALSPAC_OA.R" |
| **Two-sample MR** | Prepares data for MR | *R (4.1.1)* | **Inflammation on Cognition:**<br>"InfCog_2SMR_Prep_OA.R"<br>**Cognition on Inflammation:**<br>"InfCog_2SMR_Prep_ReverseDir_OA.R" |
| | Two-sample MR analysis | *R (4.1.1)* | **Inflammation on Cognition:**<br>"InfCog_2SMR_Analysis_OA.R"<br>**Cognition on Inflammation:**<br>"InfCog_2SMR_Analysis_Reversedir_OA.R" |
| **Downloading GWAS results** | Code to convert VCF to txt file and download online GWAS files | *Linux* | "InfCog_ConvertVCF.R" |

#### SUPPLEMENTARY FIGURES

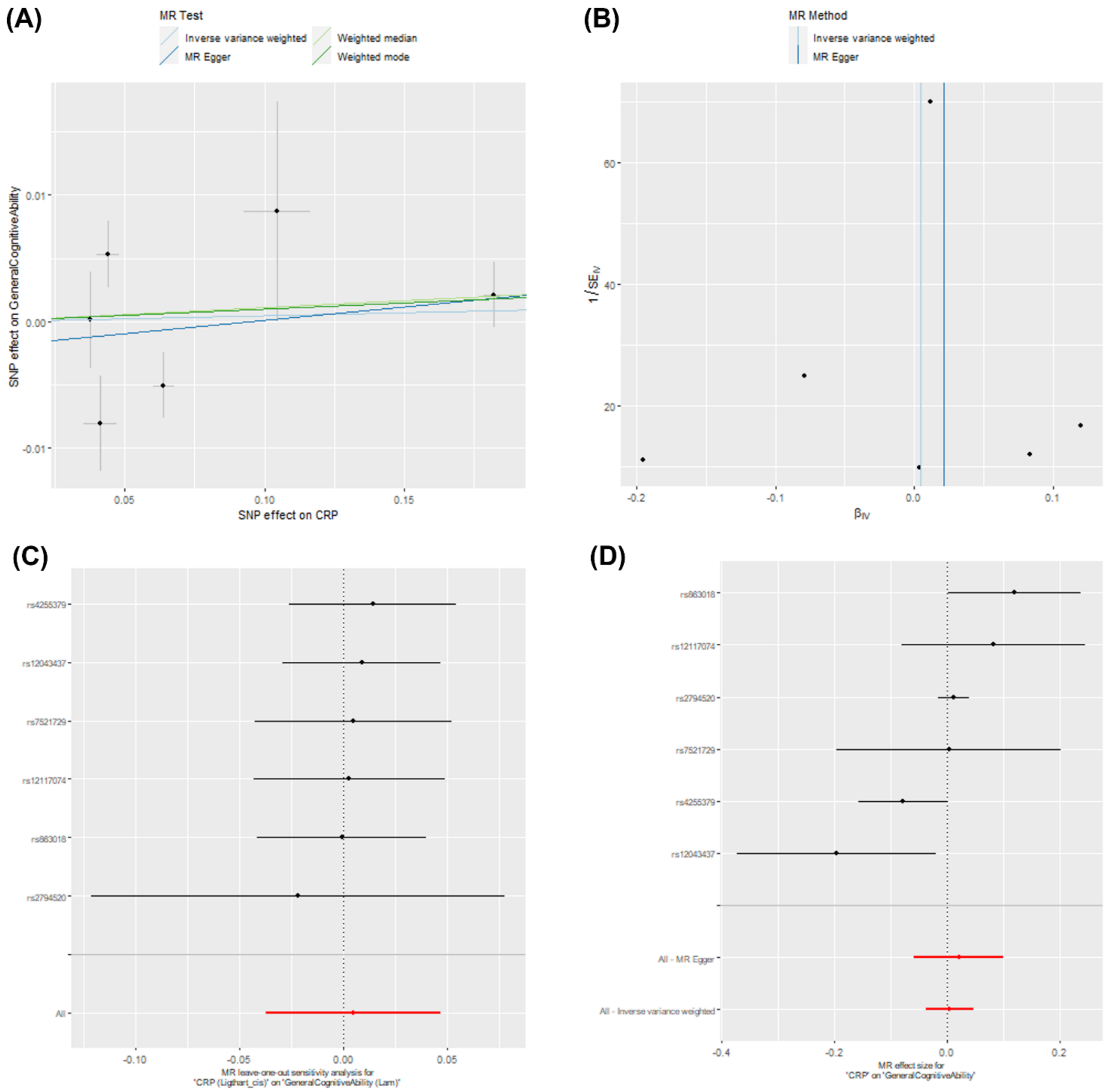

***Figure S1. Two-sample MR sensitivity plots: effect of CRP (Ligthart cis instrument) on general cognitive ability.*** Graphs include (A) scatter plot of results from four main MR methods, (B) funnel plot showing each SNP causal estimate against its precision (asymmetry may indicate directional pleiotropy), (C) leave-one-out plot showing inverse-variance weighted estimates after removing each individual SNP in turn, (D) forest plot of causal estimates for each SNP.

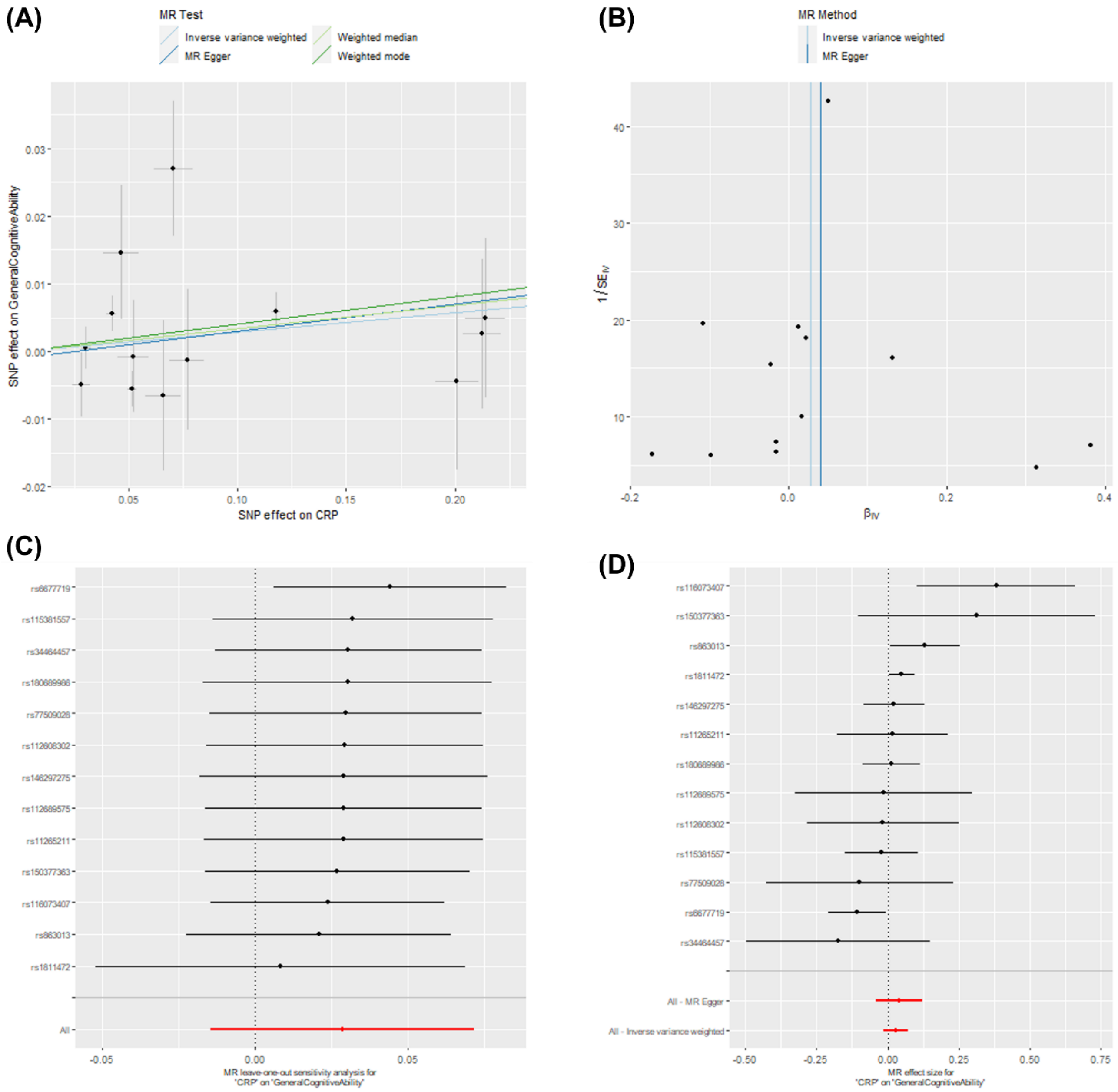

***Figure S2. Two-sample MR sensitivity plots: effect of CRP (Han cis instrument) on general cognitive ability.*** Graphs include (A) scatter plot of results from four main MR methods, (B) funnel plot showing each SNP causal estimate against its precision (asymmetry may indicate directional pleiotropy), (C) leave-one-out plot showing inverse-variance weighted estimates after removing each individual SNP in turn, (D) forest plot of causal estimates for each SNP.

(A)

(B)

(C)

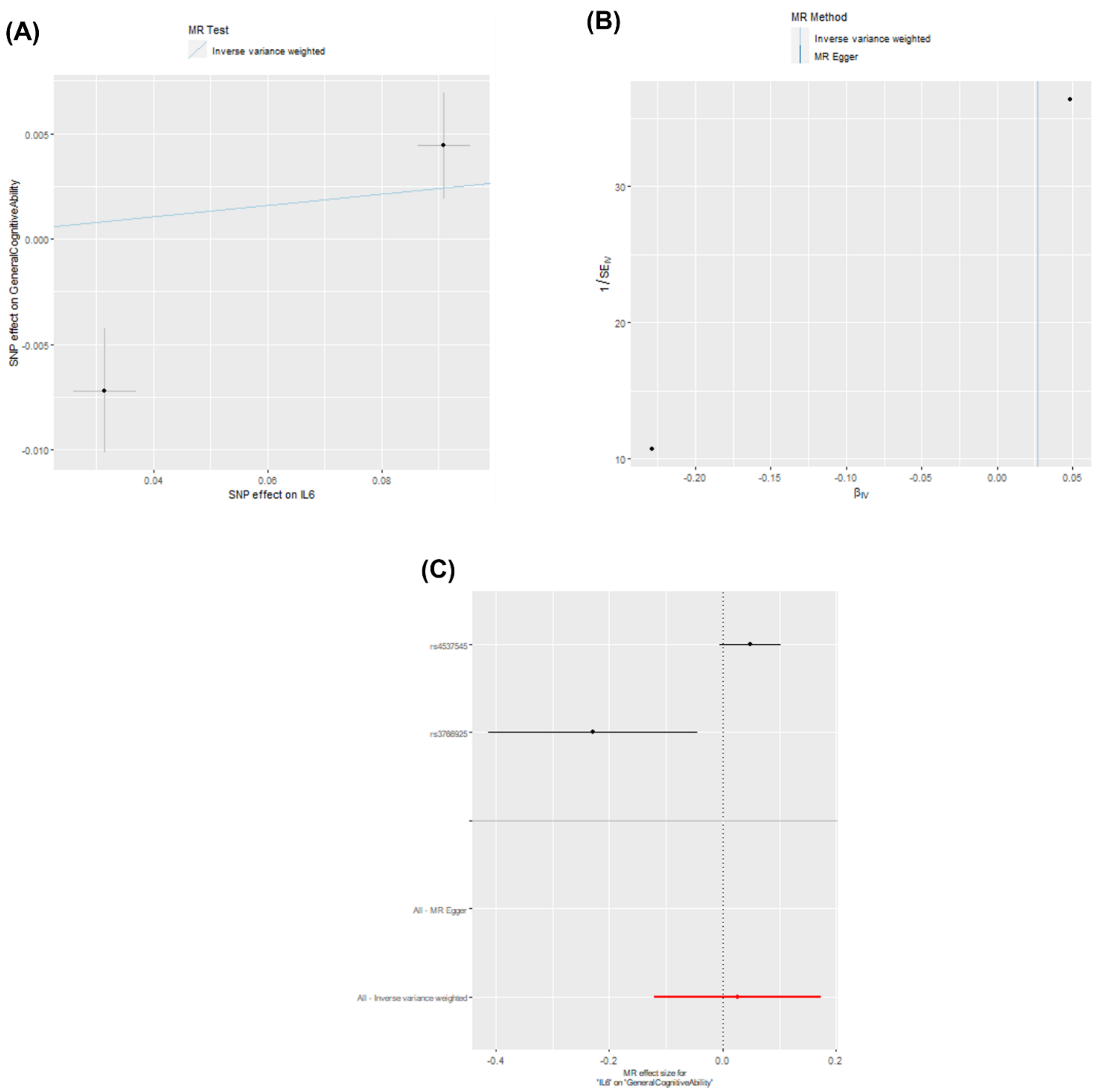

***Figure S3. Two-sample MR sensitivity plots: effect of IL-6 (Ahluwalia cis instrument) on general cognitive ability.*** Graphs include (A) scatter plot of results from inverse-variance weighted methods, (B) funnel plot showing each SNP causal estimate against its precision (asymmetry may indicate directional pleiotropy), (C) forest plot of causal estimates for each SNP. Leave-one-out plot not shown due to too few SNPs in this instrument.

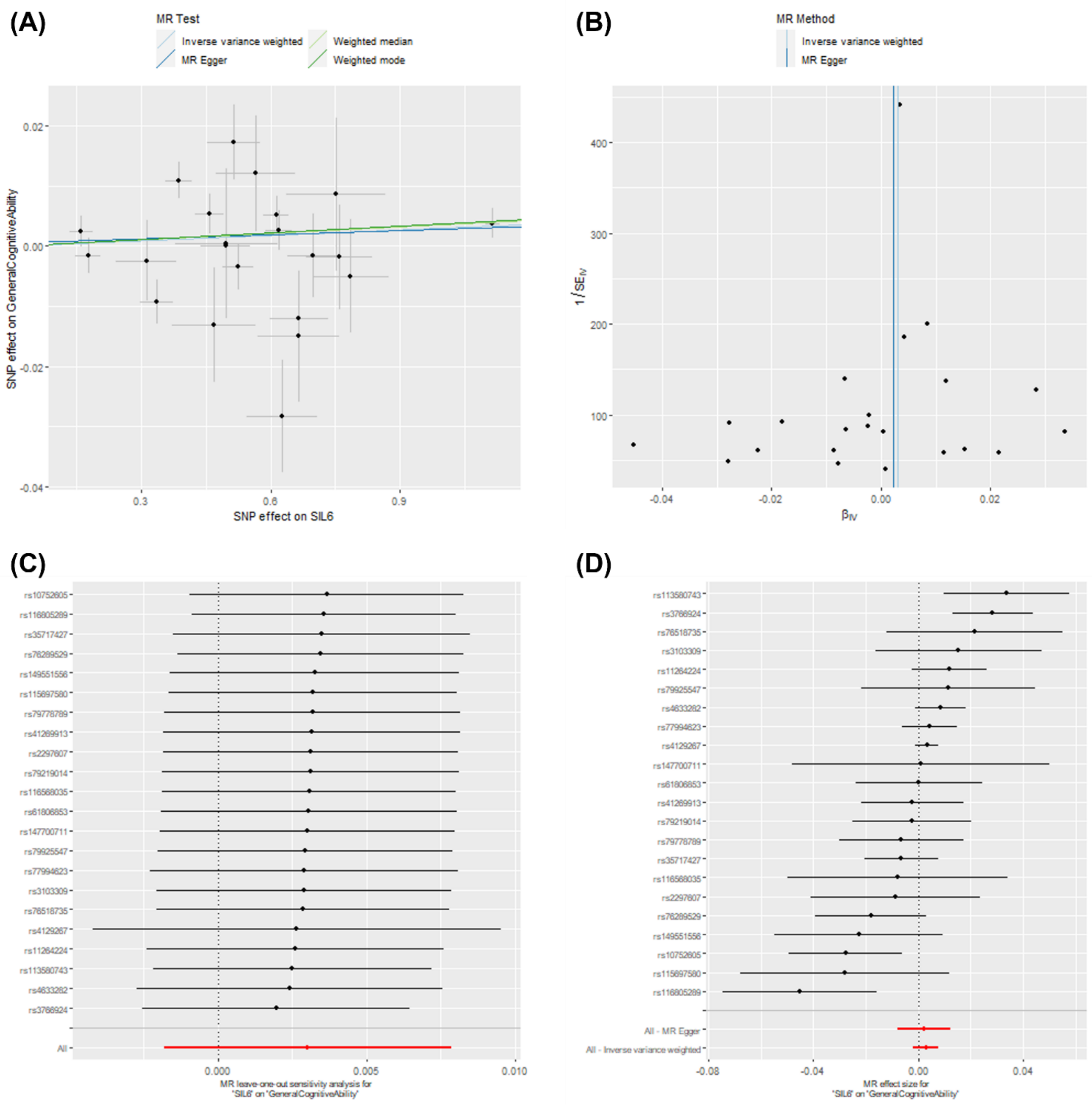

***Figure S4. Two-sample MR sensitivity plots: effect of sIL-6R (Rosa instrument) on general cognitive ability.*** Graphs include (A) scatter plot of results from four main MR methods, (B) funnel plot showing each SNP causal estimate against its precision (asymmetry may indicate directional pleiotropy), (C) leave-one-out plot showing inverse-variance weighted estimates after removing each individual SNP in turn, (D) forest plot of causal estimates for each SNP.

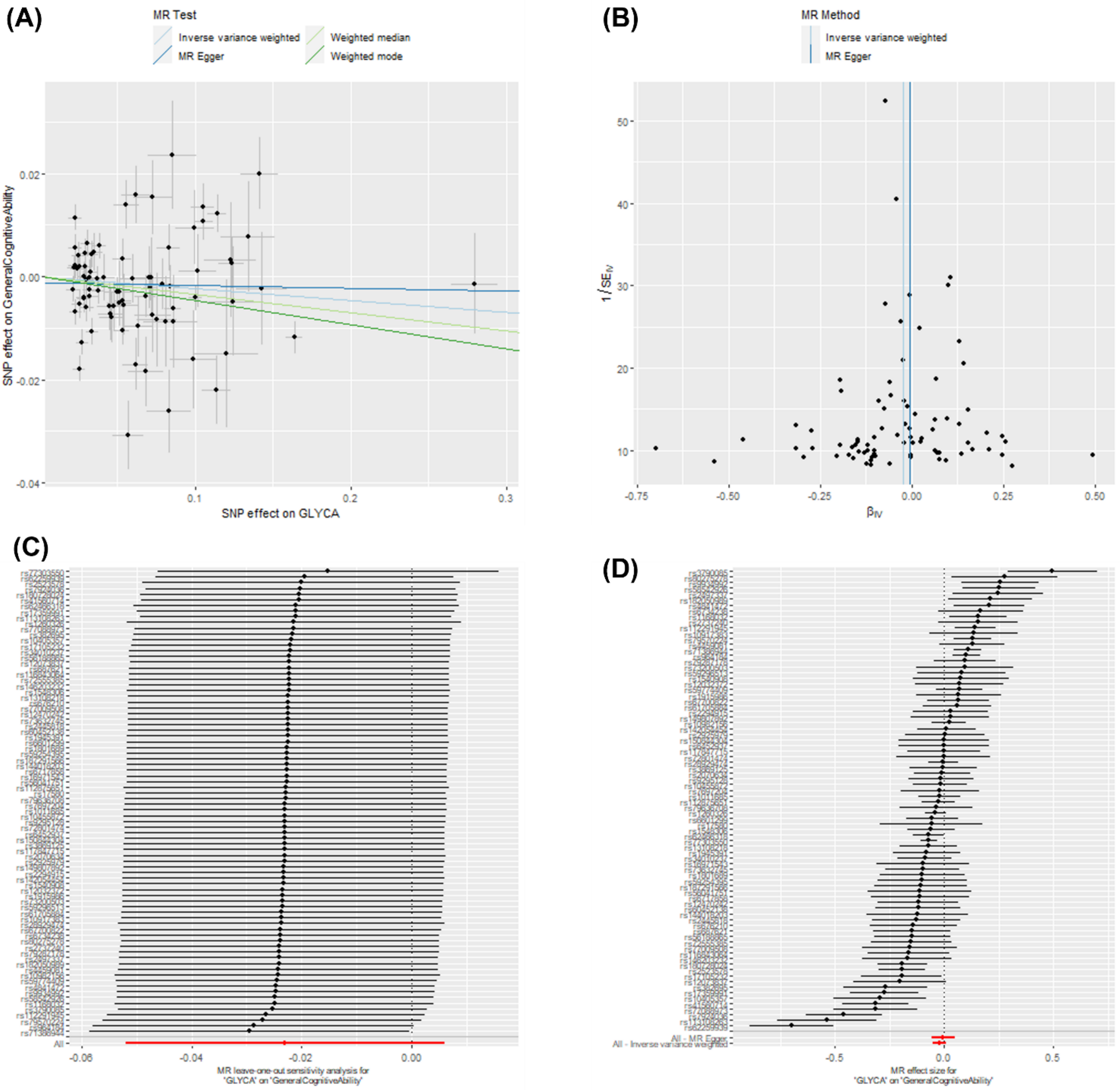

***Figure S5. Two-sample MR sensitivity plots: effect of GlycA (Borges instrument) on general cognitive ability.*** Graphs include (A) scatter plot of results from four main MR methods, (B) funnel plot showing each SNP causal estimate against its precision (asymmetry may indicate directional pleiotropy), (C) leave-one-out plot showing inverse-variance weighted estimates after removing each individual SNP in turn, (D) forest plot of causal estimates for each SNP.

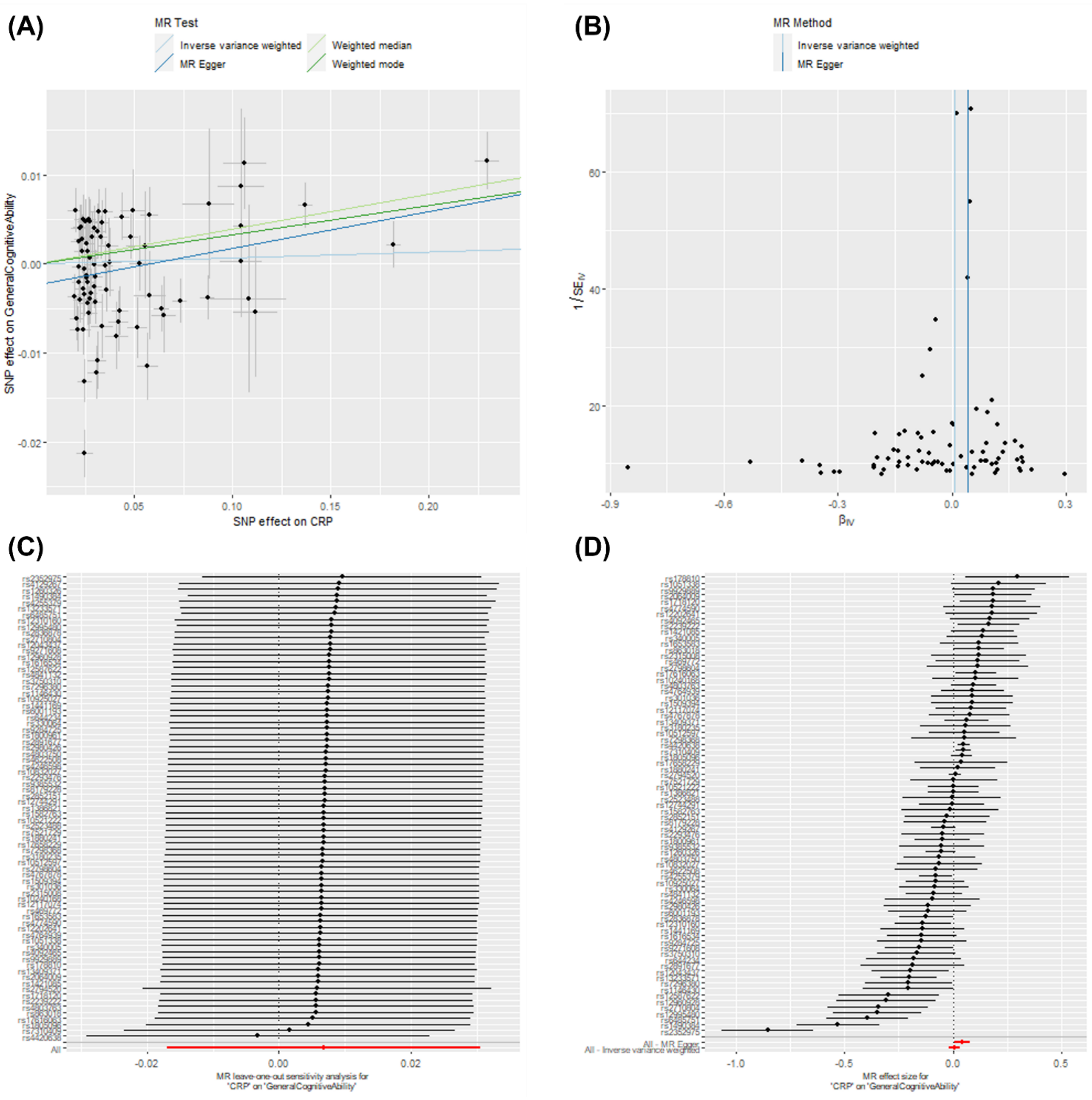

***Figure S6. Two-sample MR sensitivity plots: effect of CRP (Ligthart genome-wide instrument) on general cognitive ability.*** Graphs include (A) scatter plot of results from four main MR methods, (B) funnel plot showing each SNP causal estimate against its precision (asymmetry may indicate directional pleiotropy), (C) leave-one-out plot showing inverse-variance weighted estimates after removing each individual SNP in turn, (D) forest plot of causal estimates for each SNP.

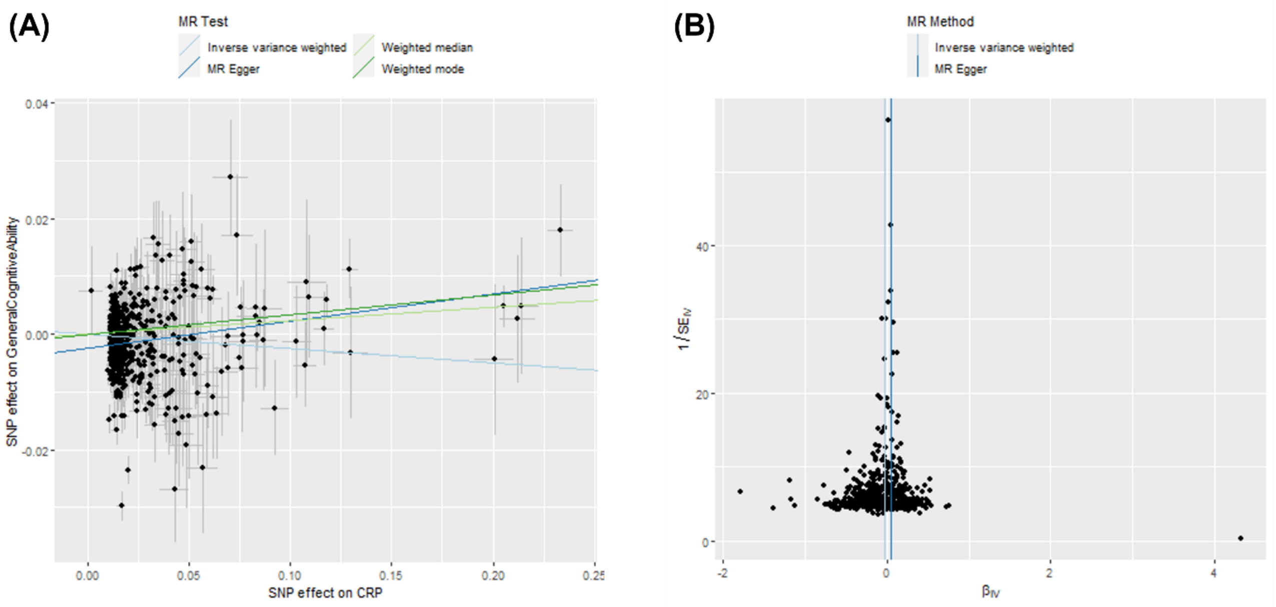

***Figure S7. Two-sample MR sensitivity plots: effect of CRP (Han genome-wide instrument) on general cognitive ability.*** Graphs include (A) scatter plot of results from four main MR methods, (B) funnel plot showing each SNP causal estimate against its precision (asymmetry may indicate directional pleiotropy). Due to a large number of SNPs being used, leave-one-out plot and forest plot are not shown due to poor visibility visualising all SNPs.

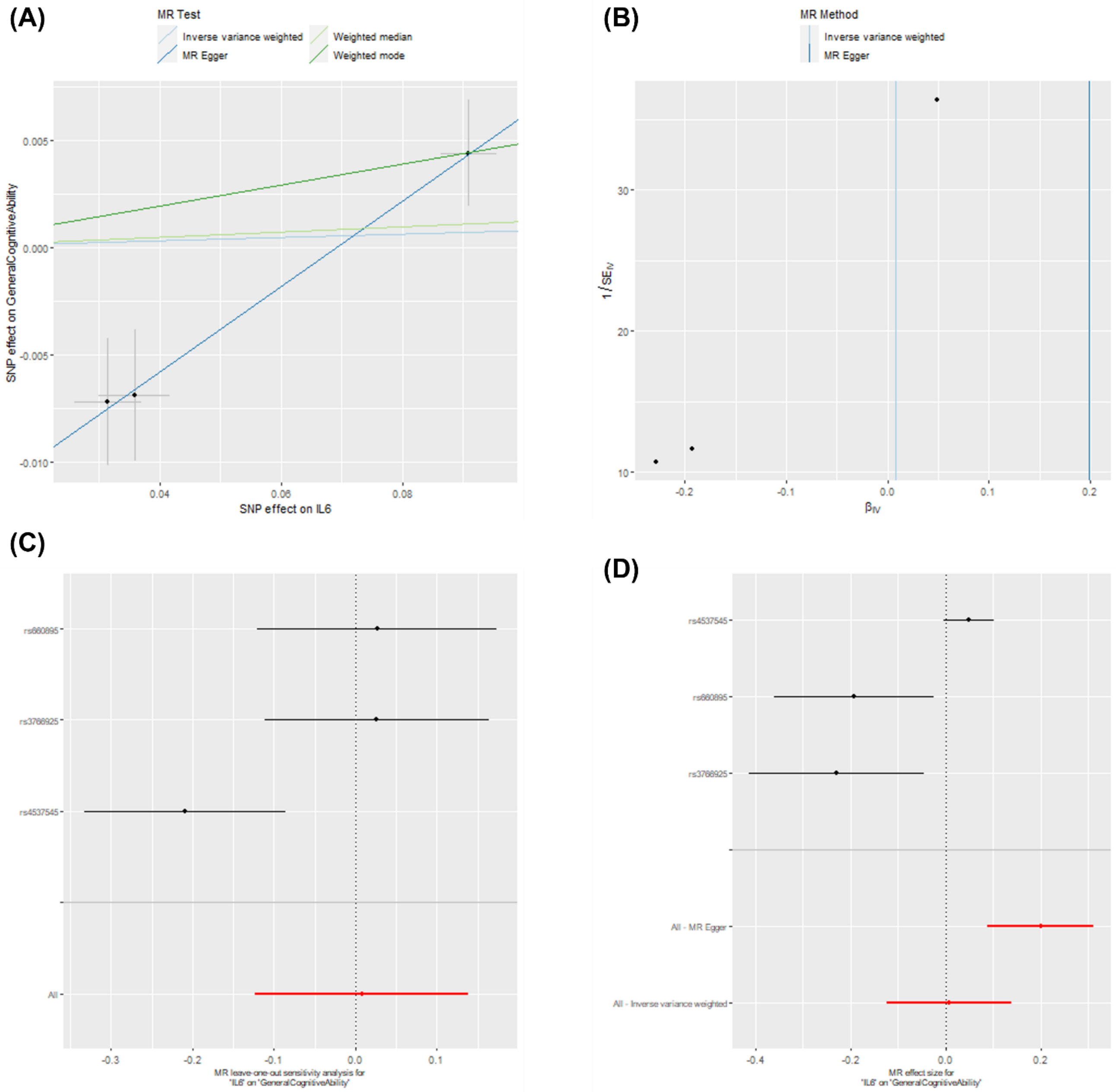

***Figure S8. Two-sample MR sensitivity plots: effect of IL-6 (Ahluwalia genome-wide instrument) on general cognitive ability.*** Graphs include (A) scatter plot of results from four main MR methods, (B) funnel plot showing each SNP causal estimate against its precision (asymmetry may indicate directional pleiotropy), (C) leave-one-out plot showing inverse-variance weighted estimates after removing each individual SNP in turn, (D) forest plot of causal estimates for each SNP.

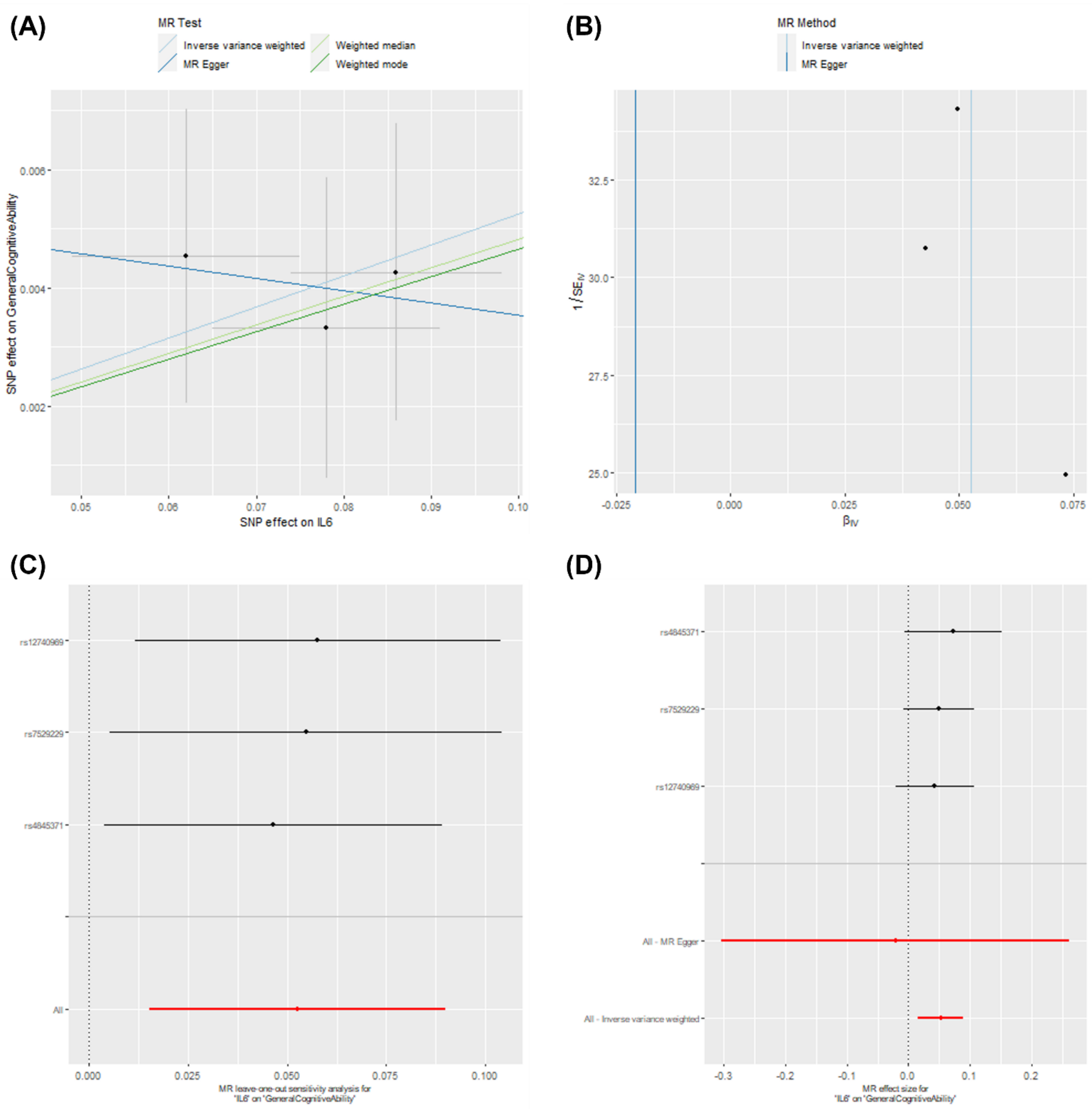

***Figure S9. Two-sample MR sensitivity plots: effect of IL-6 (Swerdlow instrument) on general cognitive ability.*** Graphs include (A) scatter plot of results from four main MR methods, (B) funnel plot showing each SNP causal estimate against its precision (asymmetry may indicate directional pleiotropy), (C) leave-one-out plot showing inverse-variance weighted estimates after removing each individual SNP in turn, (D) forest plot of causal estimates for each SNP.

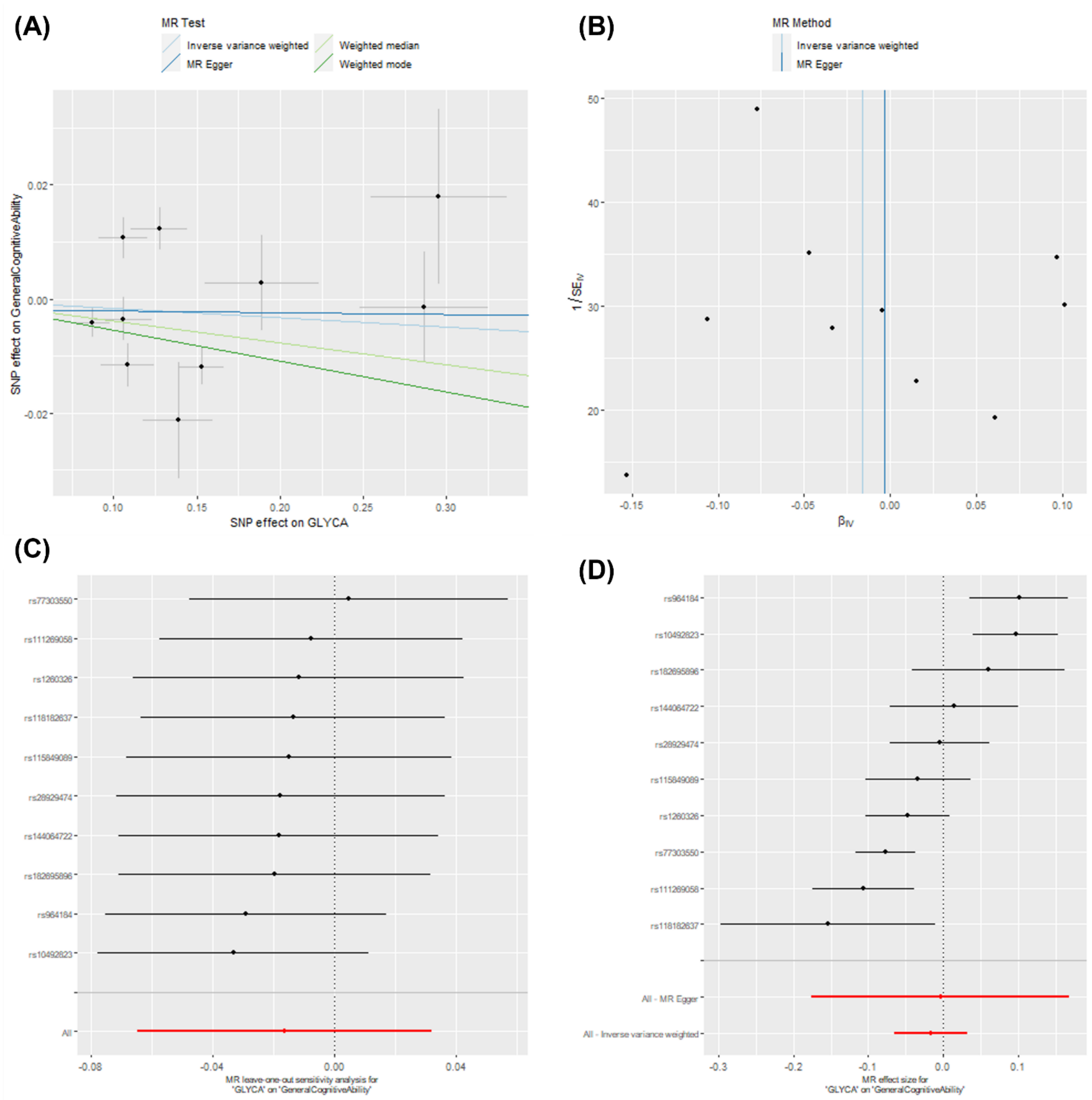

***Figure S10. Two-sample MR sensitivity plots: effect of GlycA (Kettunen instrument) on general cognitive ability.*** Graphs include (A) scatter plot of results from four main MR methods, (B) funnel plot showing each SNP causal estimate against its precision (asymmetry may indicate directional pleiotropy), (C) leave-one-out plot showing inverse-variance weighted estimates after removing each individual SNP in turn, (D) forest plot of causal estimates for each SNP.

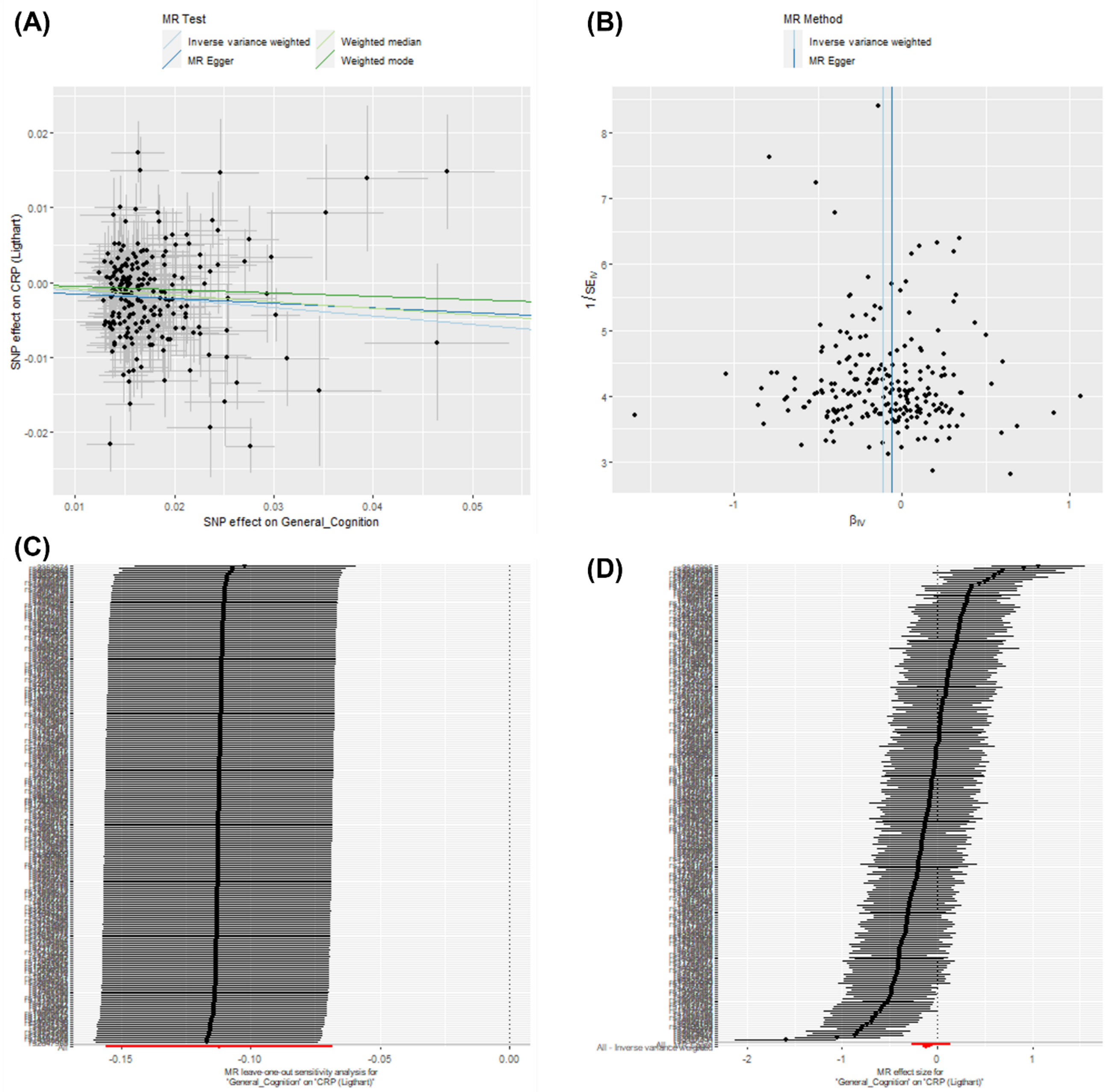

***Figure S11. Two-sample MR sensitivity plots: effect of general cognitive ability on CRP (Ligthart et al. GWAS).*** Graphs include (A) scatter plot of results from four main MR methods, (B) funnel plot showing each SNP causal estimate against its precision (asymmetry may indicate directional pleiotropy), (C) leave-one-out plot showing inverse-variance weighted estimates after removing each individual SNP in turn, (D) forest plot of causal estimates for each SNP.

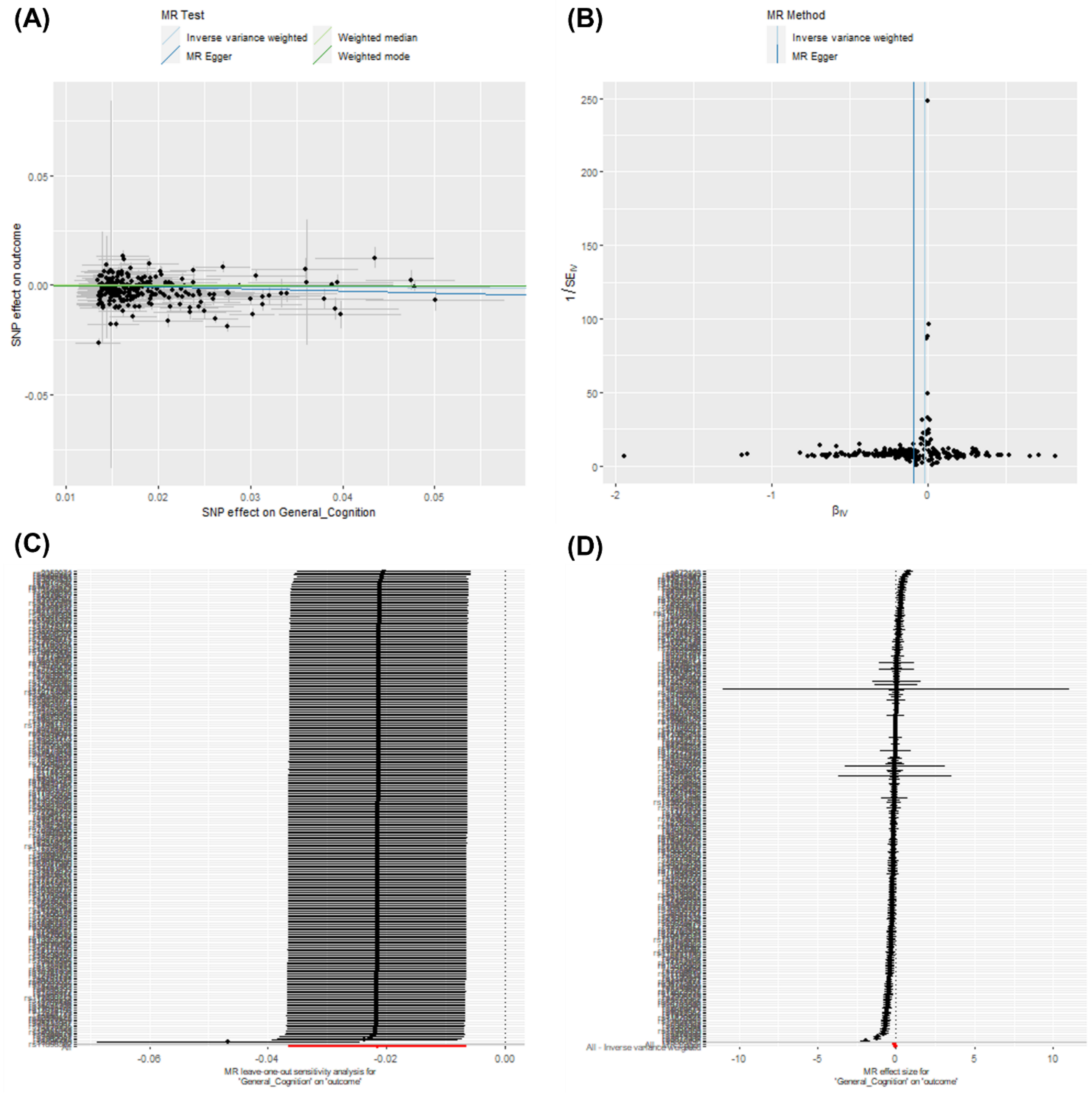

***Figure S12. Two-sample MR sensitivity plots: effect of general cognitive ability on CRP (Han et al. GWAS).*** Graphs include (A) scatter plot of results from four main MR methods, (B) funnel plot showing each SNP causal estimate against its precision (asymmetry may indicate directional pleiotropy), (C) leave-one-out plot showing inverse-variance weighted estimates after removing each individual SNP in turn, (D) forest plot of causal estimates for each SNP.

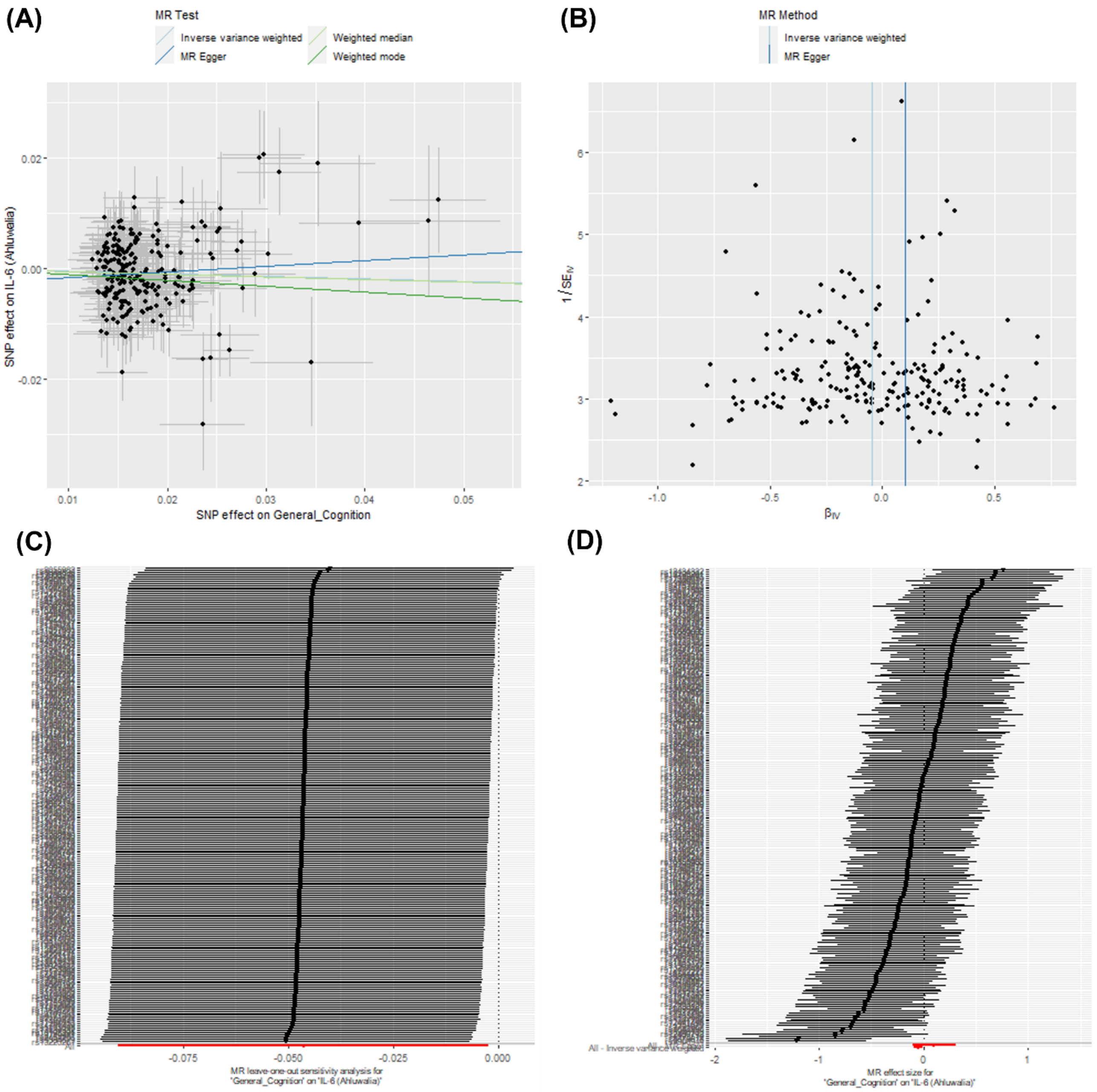

***Figure S13. Two-sample MR sensitivity plots: effect of general cognitive ability on IL-6 (Ahluwalia et al. GWAS).*** Graphs include (A) scatter plot of results from four main MR methods, (B) funnel plot showing each SNP causal estimate against its precision (asymmetry may indicate directional pleiotropy), (C) leave-one-out plot showing inverse-variance weighted estimates after removing each individual SNP in turn, (D) forest plot of causal estimates for each SNP.

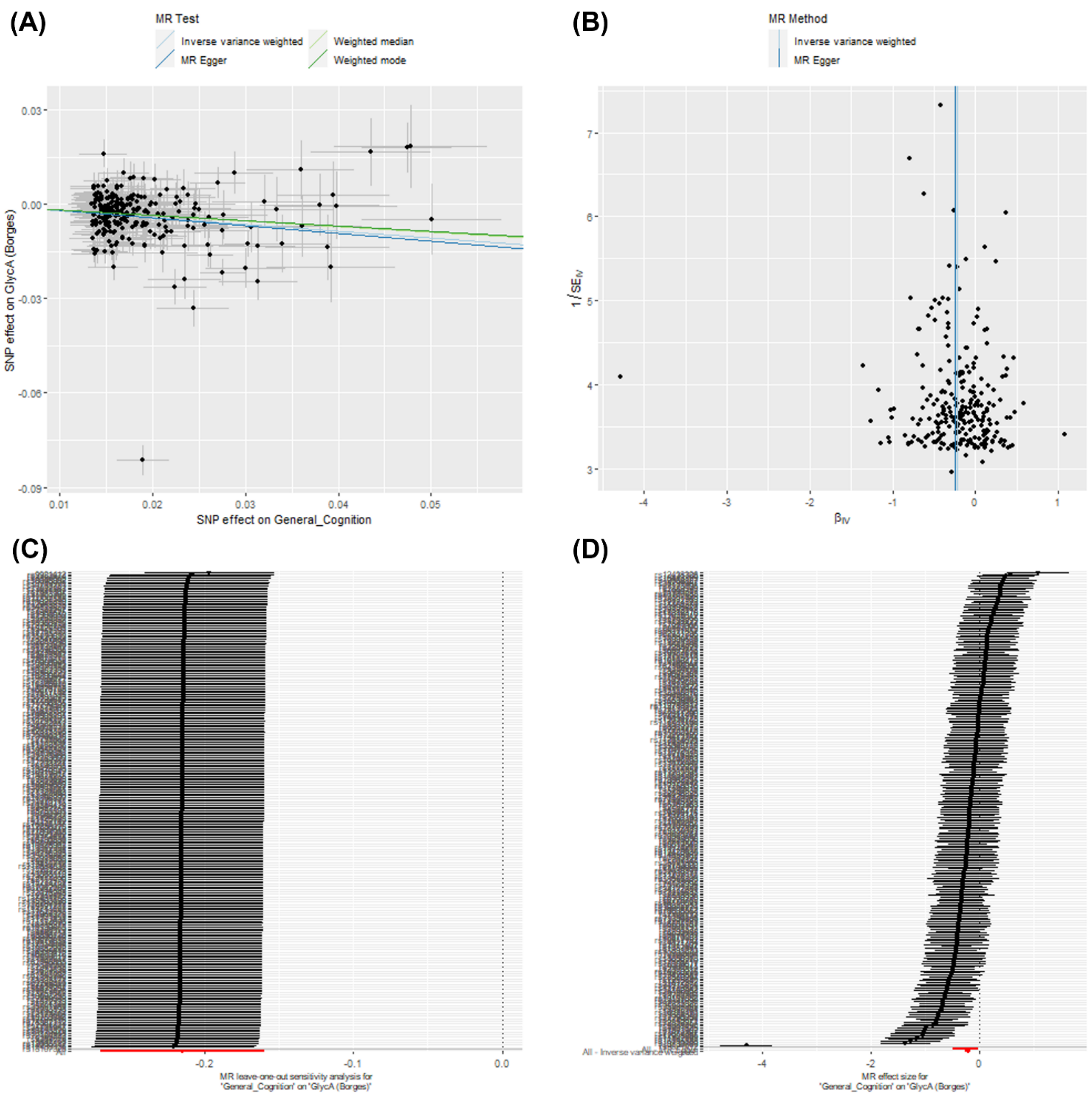

***Figure S14. Two-sample MR sensitivity plots: effect of general cognitive ability on GlycA (Borges et al. GWAS).*** Graphs include (A) scatter plot of results from four main MR methods, (B) funnel plot showing each SNP causal estimate against its precision (asymmetry may indicate directional pleiotropy), (C) leave-one-out plot showing inverse-variance weighted estimates after removing each individual SNP in turn, (D) forest plot of causal estimates for each SNP.

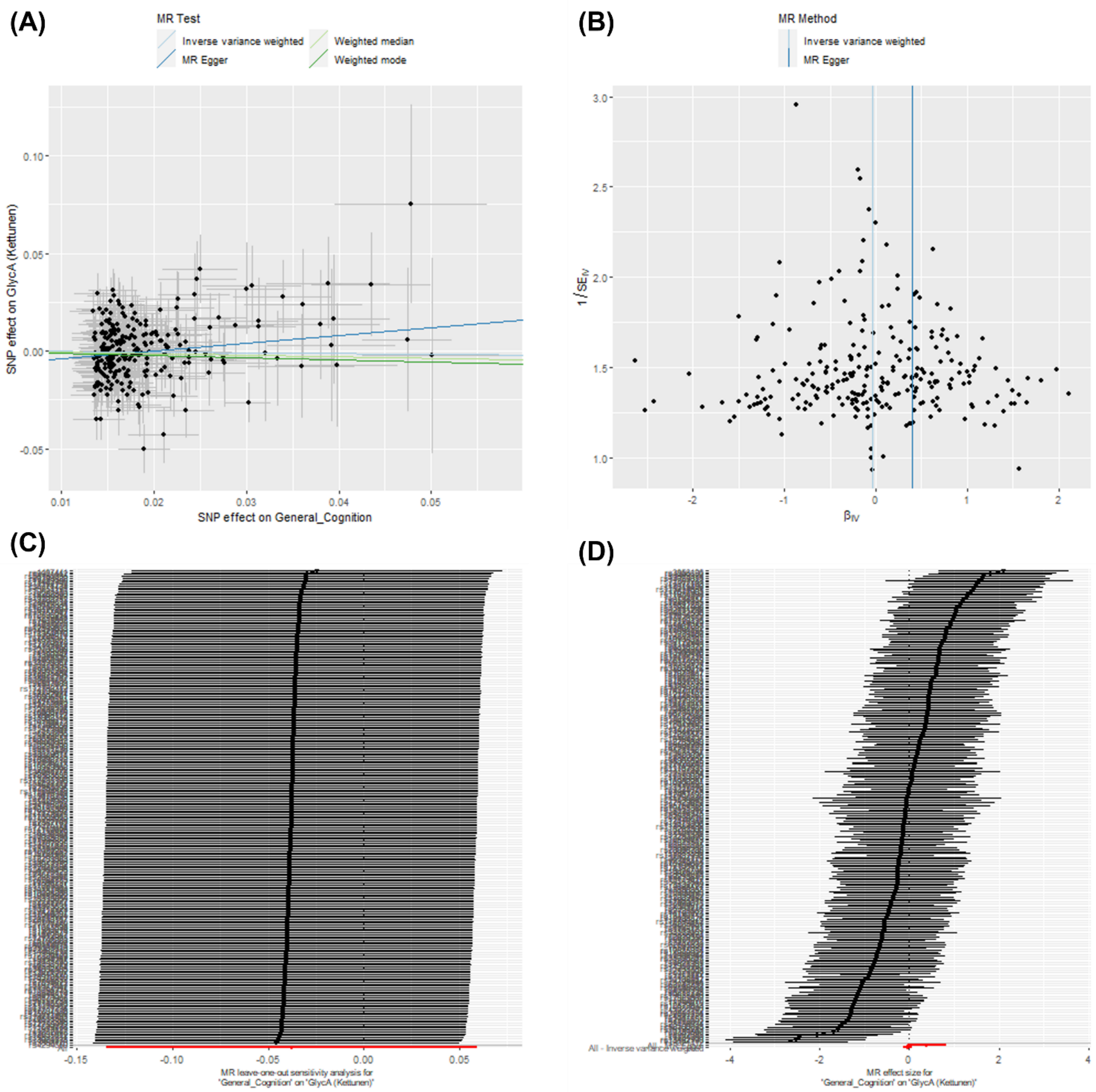

***Figure S15. Two-sample MR sensitivity plots: effect of general cognitive ability on GlycA (Kettunen et al. GWAS).*** Graphs include (A) scatter plot of results from four main MR methods, (B) funnel plot showing each SNP causal estimate against its precision (asymmetry may indicate directional pleiotropy), (C) leave-one-out plot showing inverse-variance weighted estimates after removing each individual SNP in turn, (D) forest plot of causal estimates for each SNP.

analysis. *Lancet* 379: 1214–1224.
